# Computing the daily reproduction number of COVID-19 by inverting the renewal equation using a variational technique

**DOI:** 10.1101/2020.08.01.20165142

**Authors:** Luis Alvarez, Miguel Colom, Jean-David Morel, Jean-Michel Morel

**Author notes:** L. Alvarez and J-M. Morel designed and performed research and experiments and wrote the paper. L. Alvarez implemented the method. M. Colom built the online interface and collected and processed data. J.D. Morel rewrote parts and designed the statistical analysis and presentation of the results. The authors declare no competing interests.

## Abstract

The COVID-19 pandemic has undergone frequent and rapid changes in its local and global infection rates, driven by governmental measures, or the emergence of new viral variants. The reproduction number *R*_*t*_ indicates the average number of cases generated by an infected person at time *t* and is a key indicator of the spread of an epidemic. A timely estimation of *R*_*t*_ is a crucial tool to enable governmental organizations to adapt quickly to these changes and assess the consequences of their policies. The EpiEstim method is the most widely accepted method for estimating *R*_*t*_. But it estimates *R*_*t*_ with a significant temporal delay. Here, we propose a new method, *EpiInvert*, that shows good agreement with EpiEstim, but that provides estimates of *R*_*t*_ several days in advance. We show that *R*_*t*_ can be estimated by inverting the renewal equation linking *R*_*t*_ with the observed incidence curve of new cases, *i*_*t*_. Our signal processing approach to this problem yields both *R*_*t*_ and a restored *i*_*t*_ corrected for the “weekend effect” by applying a deconvolution + denoising procedure. The implementations of the EpiInvert and EpiEstim methods are fully open-source and can be run in real-time on every country in the world, and every US state through a web interface at www.ipol.im/epiinvert.

**Significance Statement:** Based on a signal processing approach we propose a method to compute the reproduction number *R*_*t*_, the transmission potential of an epidemic over time. *R*_*t*_ is estimated by minimizing a functional that enforces: (i) the ability to produce an incidence curve *i*_*t*_ corrected of the weekly periodic bias produced by the “weekend effect”, obtained from *R*_*t*_ through a renewal equation; (ii) the regularity of *R*_*t*_. A good agreement is found between our *R*_*t*_ estimate and the one provided by the currently accepted method, EpiEstim, except our method predicts *R*_*t*_ several days closer to present. We provide the mathematical arguments for this shift. Both methods, applied every day on each country, can be compared at www.ipol.im/epiinvert.

---

The reproduction number *R*_*t*_ is a key epidemiological parameter evaluating transmission potential of a disease over time. It is defined as the average number of new infections caused by a single infected individual at time *t* in a partially susceptible population (1). *R*_*t*_ can be computed from the daily observation of the incidence curve *i*_*t*_, but requires empirical knowledge of the probability distribution Φ_*s*_ of the delay between two infections (2, 3).

There are two different models for the incidence curve and its corresponding infection delay Φ. In a theoretical model, *i*_*t*_ would represent the real daily number of new infections, and Φ_*s*_ is sometimes called *generation time* (4, 5) and represents the probability distribution of the time between infection of a primary case and infections in secondary cases. In practice, neither parameter is easily observable because the infected are rarely detected before the appearance of symptoms and tests will be negative until the virus has multiplied over several days. What is routinely recorded by health organizations is the number of *new detected, incident cases*. When dealing with this real incidence curve, Φ_*s*_ is called *serial interval* (4, 5). The serial interval is defined as the delay between the onset of symptoms in a primary case and the onset of symptoms in secondary cases (5).

*R*_*t*_ is linked to *i*_*t*_ and Φ through the *renewal equation*, first formulated for birth-death processes in a 1907 note of Alfred Lotka (6). We adopt the Nishiura et al. formulation (7, 8),

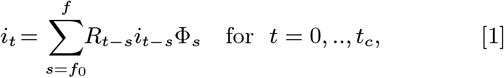

where *t*_*c*_ represents the last time at which *i*_*t*_ was available, *f*_0_ and *f* are the maximal and minimal observed times between a primary and a secondary case. The underlying epidemiological assumption of this model is that the time-varying factor *R*_*t*_ causes a constant proportional change in an individual’ s infectiousness, over the course of their entire infectious period, based on the day on which they were infected. In this case we refer to *R*_*t*_ as the case reproductive number. According to Cori et al. (5), “It is the average number of secondary cases that a case infected at time step t will eventually infect (9).”

It is important to note that secondary infections are sometimes detected before primary ones, and therefore the minimum delay *f*_0_ is generally negative (see Fig. 2). Equation [1] does not yield an explicit expression for *R*_*t*_. Yet, an easy solution can be found for the version of the renewal equation proposed in Fraser (9) (equation (9)), and Cori et al in (5),

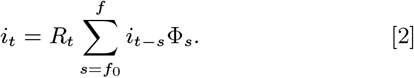

By this equation, *R*_*t*_ is derived at time *t* from the past incidence values *i*_*t*−*s*_ by a simple division, provided that *f*_0_ ≥ 0:

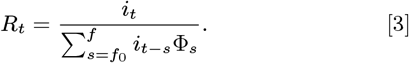

The underlying epidemiological assumption of this model is that the time-varying factor of *R*_*t*_ causes a change in the infectiousness only on the day on which transmission occurs^*^. In this case we refer to *R*_*t*_ as the instantaneous reproduction number. This *R*_*t*_ estimate, implemented by the EpiEstim software, is highly recommended in a very recent review (11) signed by representatives from ten different epidemiological labs from several continents.

EpiEstim is the standard method to compute a real-time estimation of the reproduction number, and of widespread use. In its stochastic formulation, the first member *i*_*t*_ of Equation [2] is assumed to be a Poisson variable, and the second member of this equation is interpreted as the expectation of this Poisson variable. This leads to a maximum likelihood estimation strategy to compute *R*_*t*_ (see (5, 12–15)). A detailed description of EpiEstim methods can be found in the supporting information.

Comparing Equations [2] and [1] shows that when applied with the same serial interval and case incidence curve, the second equation is derived from the first by assuming *R*_*t*_ constant on the serial interval support [*t* − *f, t* − *f*_0_]. Replacing *R*_*t*−*s*_ by *R*_*t*_ in Equation [1] indeed yields Equation [2]. A more accurate interpretation of the quotient on the right of Equation [3] would be

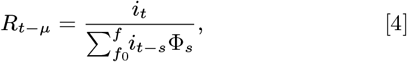

where *µ* is a central value of the probability distribution of the serial interval Φ that could be, for instance, the median or the mean. In the Ma et al. (16) estimate of the serial interval for Covid-19, we have *µ* ≃ 5.5 for the median and *µ* ≃ 6.7 for the mean. This supports the hypothesis that EpiEstim estimates *R*_*t*_ with an average delay of more than 5 days.

In practice, the way the sliding average of the incidence is calculated causes another delay. Indeed, as illustrated in Figure 1 the raw data of the incidence curve *i*_*t*_ can oscillate strongly with a seven-day period. This oscillation has little to do with the Poisson noise used in most aforementioned publications. Government statistics are affected by changes of testing and polling policies and by week-end reporting delays. These recording delays and subsequent rash corrections result in impulse noise, and a strong weekly periodic bias observable on the incidence curve (in green) on the left of figure 1.

**Fig. 1.**
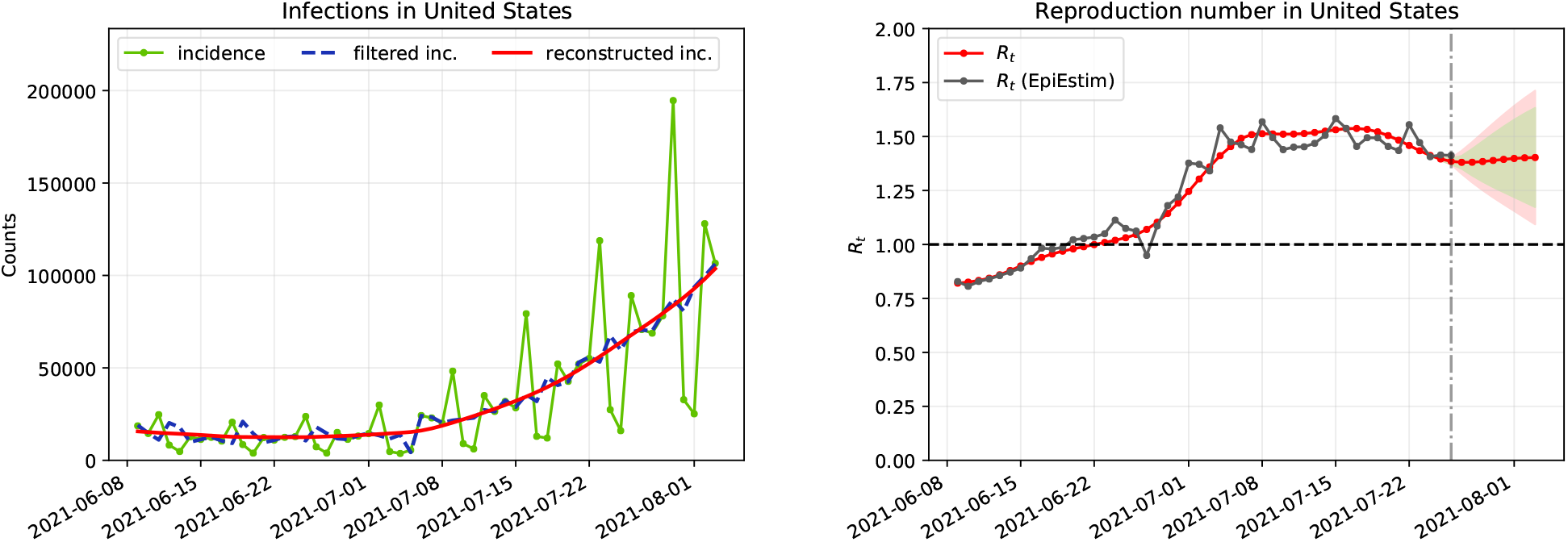
Illustration of the EpiInvert method on the USA incidence curve of new cases. On the left: in green, the raw oscillating curve of incident cases up to August 3, 2021. In blue, the incidence curve after correction of the “week-end bias”. In red, the incidence curve simulated from *R*_*t*_ after the inversion of the renewal equation. On the right: in black, *R*_*t*_, the reproduction number estimated by the current EpiEstim method, adopted by most health experts (10), shifted back eight days. Estimating its value every day guides the health policy of each country. Having *R*_*t*_ larger than 1, as it is the case for the USA on August 3, 2021 means that the pandemic is expanding. In red, the estimation of *R*_*t*_ by the EpiInvert method. This estimate, obtained by compensating the week-end bias and inverting the integral equation, has a temporal shift of about eight days with respect to EpiEstim. The shadowed areas give the 90% and 95% confidence intervals for the *R*_*t*_ estimation.

To reliably estimate the reproduction number, a regularity constraint on *R*_*t*_ is needed. Cori et al., initiators of the EpiEstim method (5), use as regularity constraint the assumption that *R*_*t*_ is locally constant in a time window of size *τ* ending at time *t* (usually *τ* = 7 days). This results in smoothing the incidence curve with a sliding mean over 7 days. This assumption has two limitations: it causes a significant resolution loss, and an additional 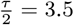 backward shift in the estimation of *R*_*t*_, given that *R*_*t*_ is assumed constant in [*t* − *τ, t*].

In summary, the computation of *R*_*t*_ by equations Eq. (1) and Eq. (2) raises three challenges:

1. The renewal equation Eq. (1) involves future values of *i*_*t*_, those for *t* + 1, …, *t* − *f*_0_.
2. Its second form Eq. (2) used by the standard method estimates *R*_*t*_ with a backward shift of about 5 days.
3. Smoothing of the week-end effect causes a 3.5 days shift backward.

These cumulative backward shifts may cause a time delay of up to 8.5 days. We shall give an experimental confirmation of such delays by two independent methods: using a simulator with synthetic ground truths, and a thorough study of the incidence curves of 55 countries. The practical meaning of this study is that the value of *R*_*t*_ computed by EpiEstim at time *t* might refer approximately to *R*_*t*−8_^†^.

Here, we address these three issues by proposing a method to invert the renewal equations Eq. (1) and Eq. (2). The inversion method developed for Eq. (1) is illustrated in Figure 1 (right), where the EpiEstim result using the renewal equation Eq. (2) (in black) is superposed with the estimate (in red) of *R*_*t*_ by EpiInvert using Eq. (1). After registering both, the black EpiEstim curve stops eight days before EpiInvert, the red curve. More generally we found, using the incidence curve of 55 countries, that the median of the temporal shift between the EpiEstim and EpiInvert *R*_*t*_ estimates using the form Eq. (1) of the renewal equation is about 8.24 days, and that the median of the RMSE approximation error between both estimates is just about 0.036.

This result is slightly surprising, given that the interpretation of *R*_*t*_ in both equations is different, and that the serial interval used in both equations also is different. In Eq. (2) the serial interval is indeed truncated to preserve the temporal causality of this equation. This excellent 0.036 fit nevertheless suggests that the EpiInvert method, applied to the renewal equation Eq. (1), is compatible with the EpiEstim method, but brings an information closer to present. This fact will be investigated experimentally in Sections 3 and 4.

The general integral equation [1] is a functional equation in *R*_*t*_. Integral equations have been previously used to estimate *R*_*t*_: in (17), the authors estimate *R*_*t*_ as the direct deconvolution of a simplified integral equation where *i*_*t*_ is expressed in terms of *R*_*t*_ and *i*_*t*_ in the past, without using the serial interval. Such inverse problems involving noise and a reproducing kernel can be resolved through the Tikhonov-Arsenin (18) variational approach involving a regularization term. This method is widely used to solve integral equations and convolutional equations (19). The solution of the equation is estimated by an energy minimization. The regularity of the solution is obtained by penalizing high values of the derivative of the solution. Our variational formulation includes the correction of the weekly periodic bias, or “weekend effect”. The standard way to deal with a weekly periodic bias is to smooth the incidence curve by a seven days sliding mean. This implicitly assumes that the periodic bias is additive. The present study supports the idea that this bias is better dealt with as multiplicative. In the variational framework, the periodic bias is therefore corrected by estimating multiplicative periodic correction factors. This is illustrated on the left graphic of Fig. 1 where the green oscillatory curve is transformed into the blue filtered curve by the same energy minimization process that also computes *R*_*t*_ (on the right in red) and reconstructs the incidence curve up to present by evaluating the renewal equation using the computed *R*_*t*_ and the filtered incidence curve (on the left, in red).

In this work we use two versions of the renewal equation formulation to compute *R*_*t*_. It is, however, possible to formulate statistical models for *R*_*t*_ that do not take into account the serial interval and the renewal equation. For instance, in (20), the author proposes to use the model:

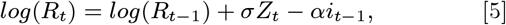

where *Z*_*t*_ is an independent and identically distributed sequence of standard normal random variables, *σ* is the dispersion of the random walk and *α* is the coefficient of drift. The model was fit to the provided incidence data by applying Bayesian inference on the parameter and state space with assumed prior distributions.

## 1. Available serial interval functions for SARS-CoV-2

As we saw, the *serial interval* in epidemiology refers to the time between successive observed cases in a chain of transmission. Du et al. in (21) define it as “the time duration between a primary case (infector) developing symptoms and secondary case (infectee) developing symptoms.”

Du et al. in (21) obtained the distribution of the serial interval by a careful inquiry on 468 pairs of patients where one was the probable cause of the infection of the other. The serial distribution Φ obtained in (21) has a significant number of cases on negative days, meaning that the infectee had developed symptoms up to *f*_0_ = 10 days before the infector. In addition to this first serial interval, we test a serial interval obtained by Nishiura et al. in (22) using 28 cases, which is approximated by a log-normal distribution, and a serial interval obtained by Ma et al. in (16) using 689 cases. As proposed by the authors this serial interval has been approximated by a shifted log-normal to take into account the cases in the negative days. In Fig. 2 we show the profile of the three serial intervals. There is good agreement of the serial intervals obtained by Du et al. (21) and Ma et al. (16)^‡^. Note that *f*_0_ = − 4 for the Ma et al. serial interval, *f*_0_ = 0 for Nishiura et al. and *f*_0_ = − 10 for Du et al. The discrete support of Φ is therefore contained in the interval [*f*_0_, *f*].

**Fig. 2.**
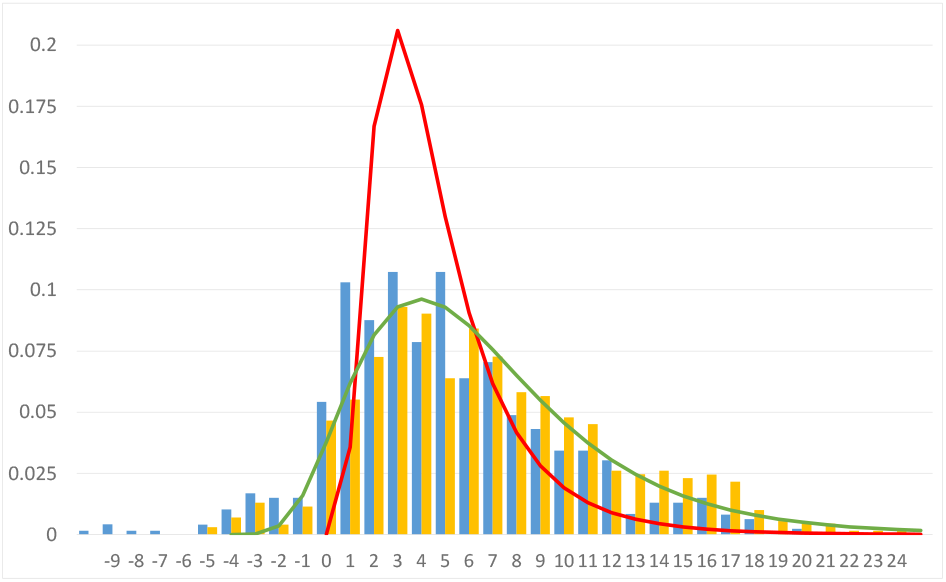
Serial intervals used in our experiments: the discrete one proposed by Du et al. in (21) (solid bars in blue), the serial interval proposed by Ma et al. (16) (solid bars inorange) and its shifted log-normal approximation (in green), finally a log-normal approximation of the serial interval proposed by Nishiura et al. in (22) (in red).

We are assuming that the serial interval profile does not change across the time. As stated in (23) (equation [10]), it is nevertheless possible to use a more general form of the renewal equation (1),

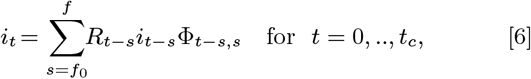

where Φ_*t*−*s,s*_ is the forward serial interval which takes into account that the onset of symptoms and transmission potential can jointly depend on the life history of a disease. The forward serial interval measures the time forward from symptom onset of an infector, obtained from a cohort of infectors that developed symptoms at the same time *t* − *s*. This more general form of the renewal equation is used in (23) to properly link the initial epidemic growth to the reproduction number *R*0. The variational approach proposed in the present work can be easily extended to compute *R*_*t*_ from *i*_*t*_ and Φ_*p,s*_, provided an estimation of Φ_*p,s*_ is available.

Transmissibility can also depend on coronavirus lineage. For instance in (24), the authors show that the SARS-CoV-2 variant B.1.1.7 has a 43 to 90% higher reproduction number than preexisting variants. It cannot be ruled out that these new variants have a different serial interval than preexisting ones.

## 2. Computing *R*_*t*_ by a variational method

As explained in the previous section, we aim at solving two versions of the renewal equation

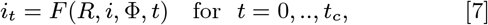

Where

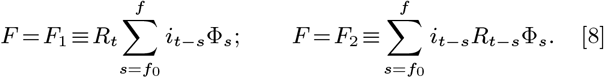

*F*_2_ corresponds to the case reproductive number formulation (equation [1]) and *F*_1_ to the instantaneous reproduction number formulation (equation [2]). Both formulations of the renewal equation are valid, and we can apply our methodology to both. As we shall see, this leads to anticipate by several days the estimate of *R*_*t*_. Equation [2] is also used in the classic Wallinga Teunis method (4), as shown in the supporting information. This last method is widely used to compute *R*_*t*_ retrospectively.

### Correcting the week-end effect

We must first formulate a compensation for the weekend effect, which in most countries is stationary, strong, and the main cause of discrepancy between *i*_*t*_ and its expected value *F* (*i, R*, Φ, *t*). To remove the weekend effect we estimate periodic multiplicative factors defined by a vector **q** = (*q*_0_, *q*_1_, *q*_2_, *q*_3_, *q*_4_, *q*_5_, *q*_6_).

The variational framework we propose to estimate *R*_*t*_ is therefore given by the minimization of the energy

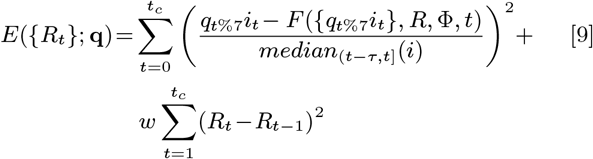

where *t*%7 denotes the remainder of the Euclidean division of *t* by 7, *t* = 0 represents the beginning of the epidemic spread and *t*_*c*_ the date of the last available incidence value.

The weekend effect has varied over the course of the pandemic. Hence, for the estimate of **q** it is better to use a time interval [*t*_*c* −_*T* + 1, *T*] where *T* is fixed i n t he experiments to *T* = 56 (8 weeks). This two months time interval is long enough to avoid overfitting and small enough to ensure that the testing policy has not changed too much. The optimization of *R*_*t*_ is instead performed through the whole time interval [0, *t*_*c*_]. The corrected value 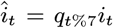 amounts to a deterministic attenuation of the weekend effect on *i*_*t*_. An obvious objection is that this correction might not be mean-preserving.

To preserve the number of accumulated cases in the period of estimation, we therefore add the constraint

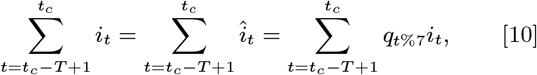

to the minimization problem [9].

In that way, the multiplication by the factor ***q***_*t*%7_ produces a redistribution of the cases *i*_*t*_ during the period of estimation, but it does not change the global amount of cases. In Equation [9], *median*_(*t* − *τ,t*]_(*i*) is the median of *i*_*t*_ in the interval (*t* − *τ, t*] used to normalize the energy with respect to the size of *i*_*t*_. In the experiments we use *τ* = 21. The first term of *E* is a data fidelity term which forces the renewal equation [7] to be satisfied as much as possible. The second term is a classic Tikhonov-Arsenin regularizer of *Rt*. As in the case of EpiEstim, this method provides a real-time estimate of *R*_*t*_ up to the date, *t*_*c*_, of the last available incidence value. Yet, in contrast with EpiEstim, this method takes advantage for *t < t*_*c*_ of the knowledge of the incidence curve 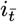 for 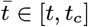.

This improves the posterior accuracy of the *R*_*t*_ estimate.

### The regularization weight

The regularization weight *w* ≥ 0 is a dimensionless constant weight fixing the balance between the data adjustment term and the regularization term.

### Boundary conditions of the variational model

Since *t* = 0 is the beginning of the epidemic spread where the virus runs free, one is led to use an estimate of *R*_0_ = *R*0 based on the basic reproduction number *R*0. (In the supporting information we present a basic estimation of *R*0 from the initial exponential growth rate of the epidemic obtained as in (25)), therefore, to solve Equation [9], we add the boundary condition *R*_0_ = *R*0. The proposed inversion model provides an estimation of *R*_*t*_ up to the date, *t*_*c*_, of the last available incidence value. Yet if *f*_0_ *<* 0, the functional [9] involves a few future values of *R*_*t*_ and *i*_*t*_ for *t*_*c ≤*_*t* _*≤*_*t*_*c* −_ *f*_0_. These values are unknown at present time *t*_*c*_. We use a basic linear regression using the last seven values of *i*_*t*_ to extrapolate the values of *i*_*t*_ beyond *t*_*c*_. We prove in the supporting information, that the boundary conditions and the choice of the extrapolation procedure have a minor influence in the estimation of *R*_*t*_ in the last days when minimizing [9].

All of the experiments described here can be reproduced with the online interface available at www.ipol.im/epiinvert. This online interface allows one to assess the performance of the method applied to the total world population and to any country and any state in the USA, with the last available data. We detail our daily sources in the supporting information.

### An empirical confidence interval for *R*_*t*_

In absence of a statistical model on the distribution of *R*_*t*_, no theoretical *a priori* confidence interval for this estimate can be given. Nevertheless, a realistic confidence interval is obtained by the following procedure: let us denote by 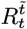 the EpiInvert estimate at time *t* using the incidence curve up to the date 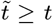. Therefore 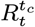 represents the final EpiInvert estimate of *R*_*t*_ using the incidence data up to the last available date *t*_*c*_. As shown below using the real and simulated data, for 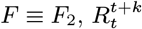 stabilizes for *k* ≥ 8. We can therefore consider 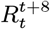 as an approximation of the reproduction number ground truth. We want to provide an empirical confidence interval 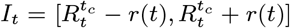 such that 95% of times 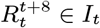 (for *t* = *t*_*c*_, *t*_*c* −_ 1, …, *t*_*c* −_ 7). To define *r*(*t*) we use, on the one hand, a measure of the variation of *R*_*t*_ in the last few days given by

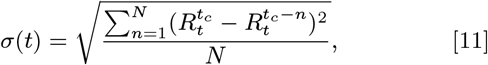

Where 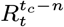 in (*t*_*c*_ − *n, t*_*c*_] are obtained by linear extrapolation. In our experiments we use *N* = 3. On the other hand, we use, supported by results obtained below for real and simulated data, that the error in the estimation of *R*_*t*_ grows linearly when *t* approaches *t*_*c*_ (the last time at which *i*_*t*_ was available). Combining *σ*(*t*) with a linear function with respect to (*t*_*c*_ −*t*) we obtain the following expression for *r*(*t*):

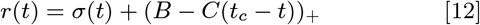

where *B* and *C* are parameters of the estimation and (*x*)_+_ ≡ *max*(0, *x*). The advantage of this empirical approach is that the estimation of the confidence interval is adapted to the variation of *R*_*t*_ in the last few days. Using 16500 experiments on real data corresponding to *R*_*t*_ estimations on 300 different values for the last used day, *t*_*c*_, in 55 countries, we obtain, in the case of *F* ≡ *F*_2_, that using *B* = 0.24 and *C* = 0.03, 95% of times 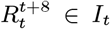 (for *t* = *t*_*c*_, *t*_*c*_ − 1, …, *t*_*c*_ − 7). In the same way for the empirical 90% confidence interval we obtain *B* = 0.16 and *C* = 0.022. If we consider now *F* ≡ *F*_1_, then it is observed that 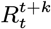 stabilizes for *k* = 3. Using it as ground truth, the obtained empirical confidence intervals for 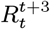 are given by *B* = 0.04 and *C* = 0.016 (in the case of 95%) and *B* = 0.02 and *C* = 0.009 (in the case of 90%) These empiric intervals are displayed for each *t* in the online algorithm www.ipol.im/epiinvert and have the aspect of fattened curves above and below *R*_*t*_.

### Efficiency measure of the weekly bias correction

We estimate the correction of the weekly periodic bias by the efficiency measure

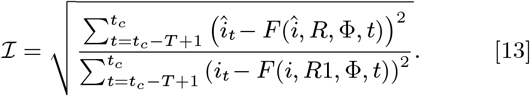

*ℐ* represents the reduction factor of the RMSE between the incidence curve and its estimate using the renewal equation after correcting the week-end bias. 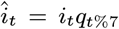 and *R* are the optimal values for the energy [9] and *R*1 denotes the *R* estimate without correction of the weekly bias. The value of *ℐ* can be used to assess whether it is worth applying the correction of the weekly periodic bias to a given country in a given time interval.

### Estimation of the temporal shift between EpiEstim and EpiIn-vert

In what follows, we will denote by 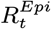 the EpiEstim estimation of the reproduction number by Cori et al. in (5), detailed in the supporting information. As we have argued above, we expect a significant temporal shift between the EpiInvert estimate of *R*_*t*_ and 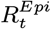, of the order of 9 days. This expectation is strongly confirmed by the experimental results, and can be checked by applying the proposed method to any country using the online interface available at www.ipol.im/epiinvert. In summary, the time shift between both methods should be a half-week (3.5 days) for *F* ≡ *F*_1_ and by Equation [4] of about *µ* + 3.5 ≃ 9 for *F* ≡ *F*_2_. This will be verified experimentally by computing the shift 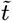 between 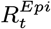 and *R*_*t*_ yielding the best RMSE between both estimates:

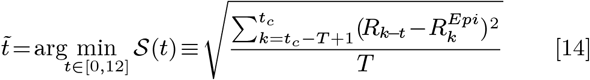

where *T* = 56 (8 weeks) and where we evaluate *R*_*k*−*t*_ for non-integer values of *k* − *t* by linear interpolation.

### Summary of the algorithm parameters and options

- choice of the serial interval: the default options are the serial intervals obtained by Ma et al. (we use the shifted log-normal approximation), Nishiura et al. and Du et al.. The users can also upload their own serial interval;
- choice of the renewal equation used, *F* ≡ *F*_1_ or *F* ≡ *F*_2_;
- Correction of the weekly periodic bias (option by default)

The regularization weight *w* is always fixed to 5, the value we obtain below, experimentally, by comparing with EpiEstim.

### Summary of the output displayed at www.ipol.im/epiinvert

First we draw two charts. In the first one we draw *R*_*t*_ and 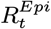 shifted back 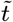 days where 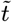 is defined in [14]. *R*_*t*_ is surrounded by a shaded area that represents the above defined empirical confidence intervals. In the second chart, we draw the initial incidence curve *i*_*t*_ in green, the incidence curve after the correction of the weekly periodic bias 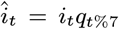 in blue, and the evaluation of the renewal equation given by 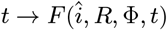 in red. For each experiment we also compute:

1. 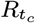: last available value of the EpiInvert *R*_*t*_ estimate.
2. 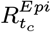: last available value of the EpiEstim estimate 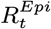.
3. 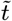: optimal shift (in days) between *R* and *R*^*Epi*^ defined in [14].
4. 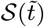: RMSE between *R* and *R*^*Epi*^ shifted back 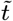 days (defined in [14]).
5. *𝒱* (*i*): variability of the original incidence curve, *i*_*t*_, given by:

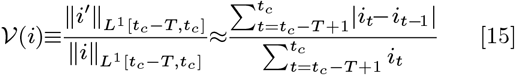
6. 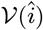: variability of the filtered incidence 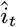 after the correction of the weekly periodic bias.
7. *ℐ*: reduction factor of the RMSE error between the incidence curve and its estimate using the renewal equation after the correction of the weekly periodic bias (defined in [13]).
8. **q** = (*q*_0_, .., *q*_6_): the correction coefficients of the weekly periodic bias (*q*_6_ corresponds to the *t*_*c*_, the last time at which *i*_*t*_ was available).

## 3. Results on incidence curves from 55 countries

To estimate a reference value for the regularization parameter *w* we used the incidence data up to July 17, 2021 for the 55 countries showing the larger number of cases. For each country, we performed 30 experiments. Starting with the incidence data up to July 17, in each experiment we removed the last 10 days from the incidence data used in the previous experiment. In that way we got a large variety of real epidemic scenarios. We optimized the 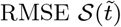 between *R*_*t*_ and 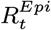 shifted back 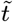 days (defined in [14]). This optimization was performed with respect to *w* and 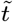. The goal was to fix *w*, the only parameter of the method, so that the result of EpiInvert is as close as possible to EpiEstim in the days where both methods predict *R*_*t*_. The second goal of this optimization was to estimate the effective time shift 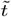 between both methods.

In Fig. 3 we show the box plot of the distribution of *w* for *F*_1_ and *F*_2_ when *w* was optimized independently for each experiment to minimize the average error over 56 days between the EpiEstim and the EpiInvert methods. The median of the distribution of *w* is 5 for *F*_1_ and *F*_2_ which indicated that a common value of *w* could be fixed for all countries. Here and in all figures to follow, each dot represents the average of all experimental results associated to a country.

**Fig. 3.**
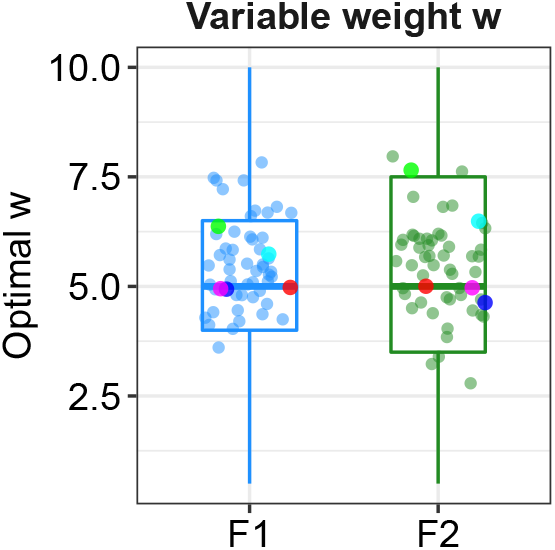
Distribution of *w* for *F*_1_ and *F*_2_ when the regularization weight *w* and th delay 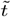 are optimized independently for each country to minimize the average error 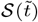 between the EpiEstim and the EpiInvert methods on a time lapse of 56 days. France in blue, Japan in green, Peru in cyan, South Africa in magenta, USA in red.

In Fig 4, we show, for the versions *F*_1_ and *F*_2_ of the renewal equation, the average error 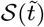 over 56 consecutive days of the error between the EpiEstim and the EpiInvert estimates of *R*_*t*_ for each country. The median of the overall average error is 0.025 for *F*_1_ and 0.034 for *F*_2_.

**Fig. 4.**
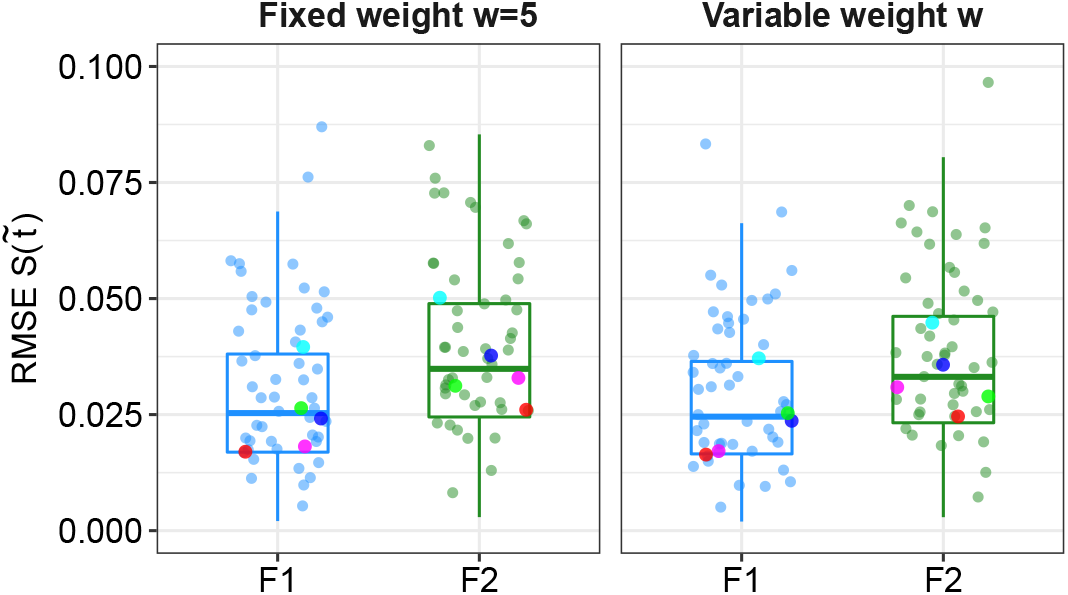
Average error 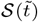 between the EpiEstim and the EpiInvert estimates of *R*_*t*_ for each country. On the left *w* is fixed and on the right it is the optimal weight per country. France in blue, Japan in green, Peru in cyan, South Africa in magenta, USA in red.

As shown in Fig. 4, the agreement between *R*_*t*_ and 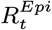 shifted back by the optimal delay 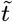 is overwhelming. As is apparent by comparing the box plots on the left and right, the increase of the error 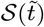 was insignificant when fixing *w* = 5 for all countries (“fixed weight”) instead of optimizing jointly on *w* and 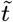 for all countries (“variable weight”). In all experiments, we therefore fixed the value of *w* to 5 for all countries. Once fixed, we optimized again 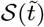 with respect to 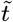.

In the box plot of Fig. 5 we show, for the versions *F*_1_ and *F*_2_ of the renewal equation, the optimal time shift 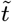 obtained by minimizing the mean error 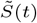 over 56 days between the EpiEstim and the EpiInvert estimates of *R*_*t*_ for each country. As is apparent by comparing the box plots on the left and right, there is almost no change on 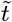 when fixing *w* for all countries (“fixed weight”) instead of optimizing jointly on *w* and 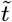 for all countries. We obtain respectively 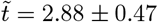 for variable *w* and 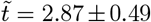 for *F*_1_ with fixed *w*, and similarly for *F*_2_: 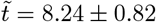 and 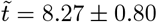. These results are in good agreement with the discussion about the EpiEstim method we have presented above, which led to predict a time delay of 3.5 days for *F* ≡ *F*_1_ and more than 8 days for *F*≡ *F*_2_. The difference between the predicted time delay and the observed one therefore is about 0.5 days. This is easily explained by the regularization term in Equation [9], which forces *R*_*t*_ to resemble *R*_*t*−1_. In summary, these experiments show that EpiEstim predicts at time *t* a value *R*_*t*_ which corresponds to day *t* − 8.5 or *t* − 3.5, and that EpiInvert predicts at time *t* a value *R*_*t*_ which corresponds to day *t* − 0.5.

**Fig. 5.**
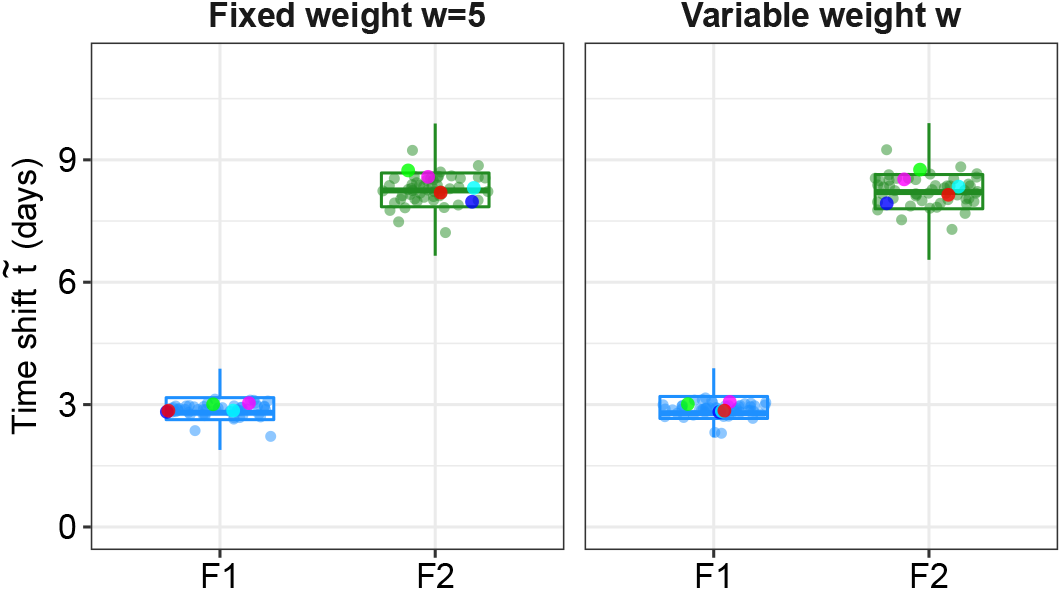
Optimal time shift 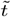 obtained by minimizing the mean error 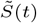 over 56 days between the EpiEstim and the EpiInvert estimates of *R*_*t*_ for each country. The time shift is, as predicted by our theoretical analysis, close to 3 days for *F*_1_ and slightly above 8 days for *F*_2_. On the left *w* is fixed and on the right it is the optimal weight per country. France in blue, Japan in green, Peru in cyan, South Africa in magenta, USA in red.

We now explore the internal coherence of the EpiInvert predictions. Let us denote by 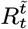 the EpiInvert estimate at time *t* using the incidence curve up to the date 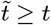. Since the estimate of EpiInvert at each day evolves with the knowledge of the incidence in later days, when 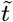 increases, the estimation 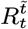 becomes more accurate and, as shown later using simulated data, we can consider that 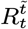 stabilizes and approaches the final estimation when 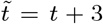 for *F* ≡ *F*_1_ and 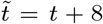 for *F* ≡ *F*_2_. Fig. 6 gives a box plot of the distributions of the internal relative error between the EpiInvert estimations depending on the prediction day *k*.

**Fig. 6.**
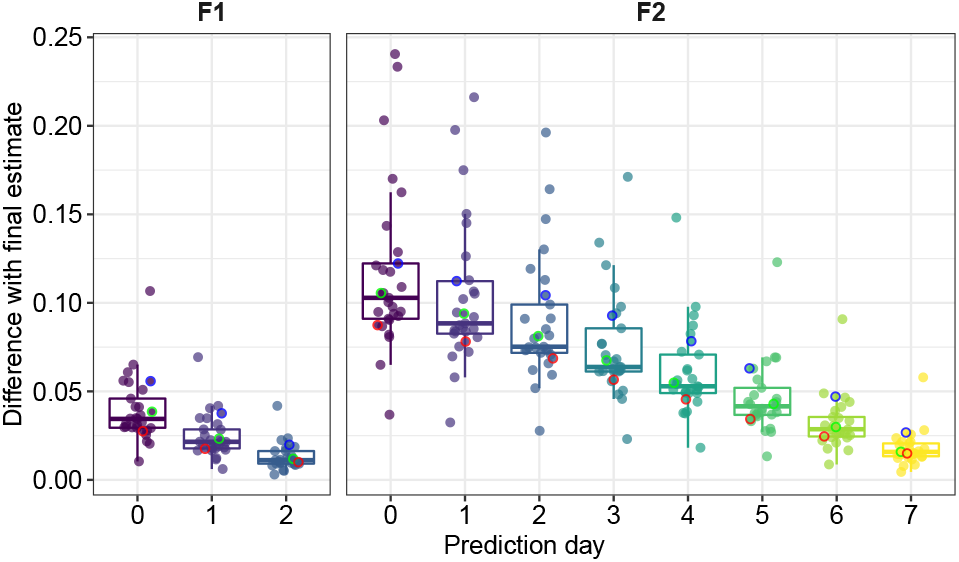
Internal relative error between the EpiInvert estimations depending on the prediction day *k*. Each dot represents the average value on 300 experiments performed on one country for different values of *t*. On the left, for *F* ≡ *F*_1_, we compare for *k* = 0, 1, 2, the relative errors 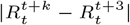. On the right, for *F* ≡ *F*_2_, we compare, in the same way, 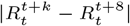 for *k* = 0, .., 7. For *F* ≡ *F*_2_, we see that 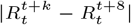 goes down almost linearly with respect to *k*. France in blue, Japan in green, Peru in black, South Africa in magenta, USA in red. The robustness of the prediction is positively affected by incidence numbers.

Fig. 7 shows, for each prediction day *k* = 0, 1, .., the linear regression of the internal relative error between the EpiInvert estimations, viewed as a function of the mean incidence of the country. These regression lines are clearly decreasing, which means that a higher incidence favors a better estimate of *R*_*t*_. Last but not least, we evaluate the reduction obtained on the “week-end effect”. Fig. 8 shows a regression plot of the RMSE reduction factor *ℐ* (see [13]) obtained by applying correcting coefficients to reduce the “week-end effect”. This reduction decreases from about 0.7 to 0.4, the plots being ordered in increasing order of average incidence. This indicates that higher incidences lead to a more regular 7 days periodicity of the week-end effect. In https://ctim.ulpgc.es/covid19/BoxPlots/ Fig. 6, 7 and 8 are presented in interactive mode with tooltip detailed statistics on each country.

**Fig. 7.**
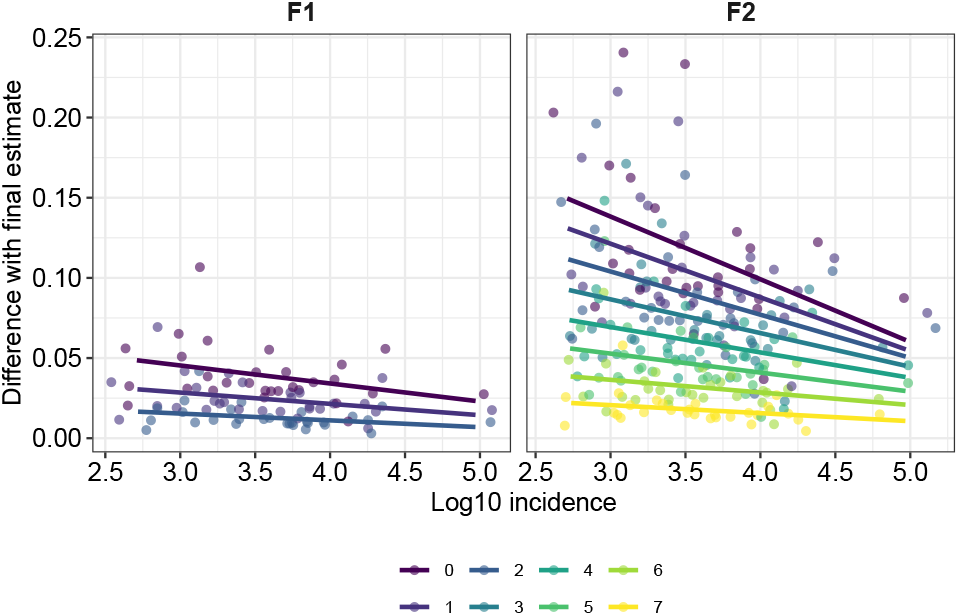
Linear regression of the internal relative error between the EpiInvert estimation as a function of the mean incidence. The regression lines are clearly decreasing, which means that a higher incidence favors a better estimate of *R*_*t*_.

**Fig. 8.**
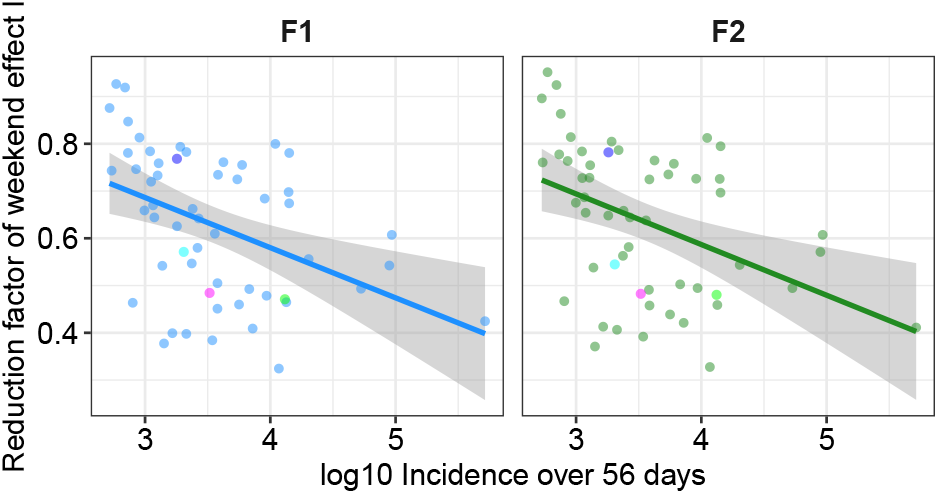
Reduction factor *ℐ* (see [13]) obtained by applying correcting coefficients to reduce the “weed end effect”. This reduction decreases from about 0.7 to less than 0.4. The plots are ordered in increasing order of average incidence.

## 4. Validation on epidemic simulations

To evaluate the accuracy of the proposed technique, we used simulated data where the ground truth for *R*_*t*_ (that we denote by 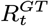) is similar to the one proposed in Gostic et al. (11). This ground truth simulates the impact of a strict lockdown at the beginning of the epidemic spread. Initially, 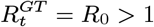, then, a strict lockdown is implemented at time *t* = 0 and 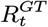 becomes *R*_*i*_ *<* 1. After *t*′ days (the lockdown duration) the social-distancing measures start relaxing to keep the *R*_*t*_ value stabilized around 1. The parameters to define 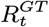 are *R*_0_, *R*_*i*_, *t*′ and *s*, which determines the slope of the transitions between *R*_0_ and *R*_*i*_ and between *R*_*i*_ and 1. the larger *s*, the sharper the transition. For a technical description of the definition of 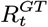, see the supporting information. The ground truth of the incidence curve, that we denote by 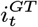, is computed from the renewal equation using 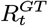 as reproduction number. Since the ground truth of the incidence curve is defined up to the multiplication by a constant factor, the simulator allows users to tune the additional parameter *i*_*max*_, which represents the maximum value of the incidence curve in the whole period. We simulated the observed incidence curve *i*_*t*_ assuming that 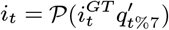 follows a Poisson distribution of mean 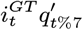 where 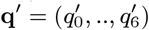 is the vector of the weekly bias correction factors.

To simulate the weekly bias the simulator proposes 19 options of real bias correction factors, pre-estimated using the incidence curve of 19 countries. Note that, in agreement with the Poisson model, the weekly bias is applied first on the deterministic incidence curve. It is followed by the Poisson simulation, which takes this biased deterministic value as parameter. The simulator finally uses EpiInvert and EpiEstim to compute *R*_*t*_ from the biased Poisson process realization *i*_*t*_. An online implementation of this simulator is available at www.ipol.im/episim.

A statistical analysis of the results was performed on 4800 simulations, obtained by varying regularly the parameters *R*_0_ ∈ [1.5, 2], *R*_*i*_ ∈ [0.5, 0.8], *s* ∈ [0.1, 2.], *i*_*max*_ ∈ [1000, 30000], *t*′ = 28, pre-estimated weekly bias from 19 countries, and the extra option of not applying weekly bias. The regularization parameter was fixed to *w* = 5, which is the optimal value obtained with real data. In the case of *F* ≡ *F*_1_, we compared the ground truth 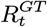 with 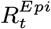 (the EpiEstim estimation), 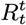 (the EpiInvert estimation using *i*_*t*_ up to the time *t*), 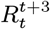 (the EpiInvert estimation at time *t* using *i*_*t*_ up to the time *t* + 3), and the final estimate 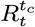. In the simulator, *t*_*c*_ *> t* + 8 is the last day used in the simulation, which depends on the lockdown duration. In the case of *F* ≡ *F*_2_, we compared the ground truth with 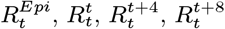 and 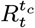.

Fig. 10 shows a thorough comparison on a lockdown scenario of the results of the *R*_*t*_ estimation methods. These simulations confirm the theoretically anticipated time delays between the various considered estimates of *R*_*t*_. Contrarily to EpiEstim, EpiInvert updates the estimated values of *R*_*t*_ when days pass by. This estimate of *R*_*t*_ obtained *k* days later, denoted by 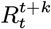, stabilizes near the (blue) ground truth 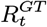 for *k* = 8 (B, red) with the F2 model, and for *k* = 3 for the F1 model (A, red). Indeed it uses, for each *t*, the incidence values up to 8 days (resp. 3 days) later. Nevertheless, for the F1 model, the timely estimate 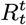 (A, orange) is very close to the ground truth 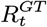(A, blue) and much closer to it than the EpiEstim estimate (A, black). For the F2 model, the timely estimate 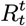 (B, orange) is not that close to 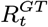. Indeed, the estimation uses the values of *i*_*t*_ up to time *t*, so there is only partial information to compute the reproduction number, which still depends on the future values of *i*_*t*_. Yet, the 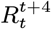 estimate (B, magenta), which is delayed by only 4 days, is considerably closer to the ground truth (in blue), and 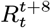 is still closer. Observe that EpiEstim (in black) provides by far the worst estimation of 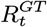.

**Fig. 9.**
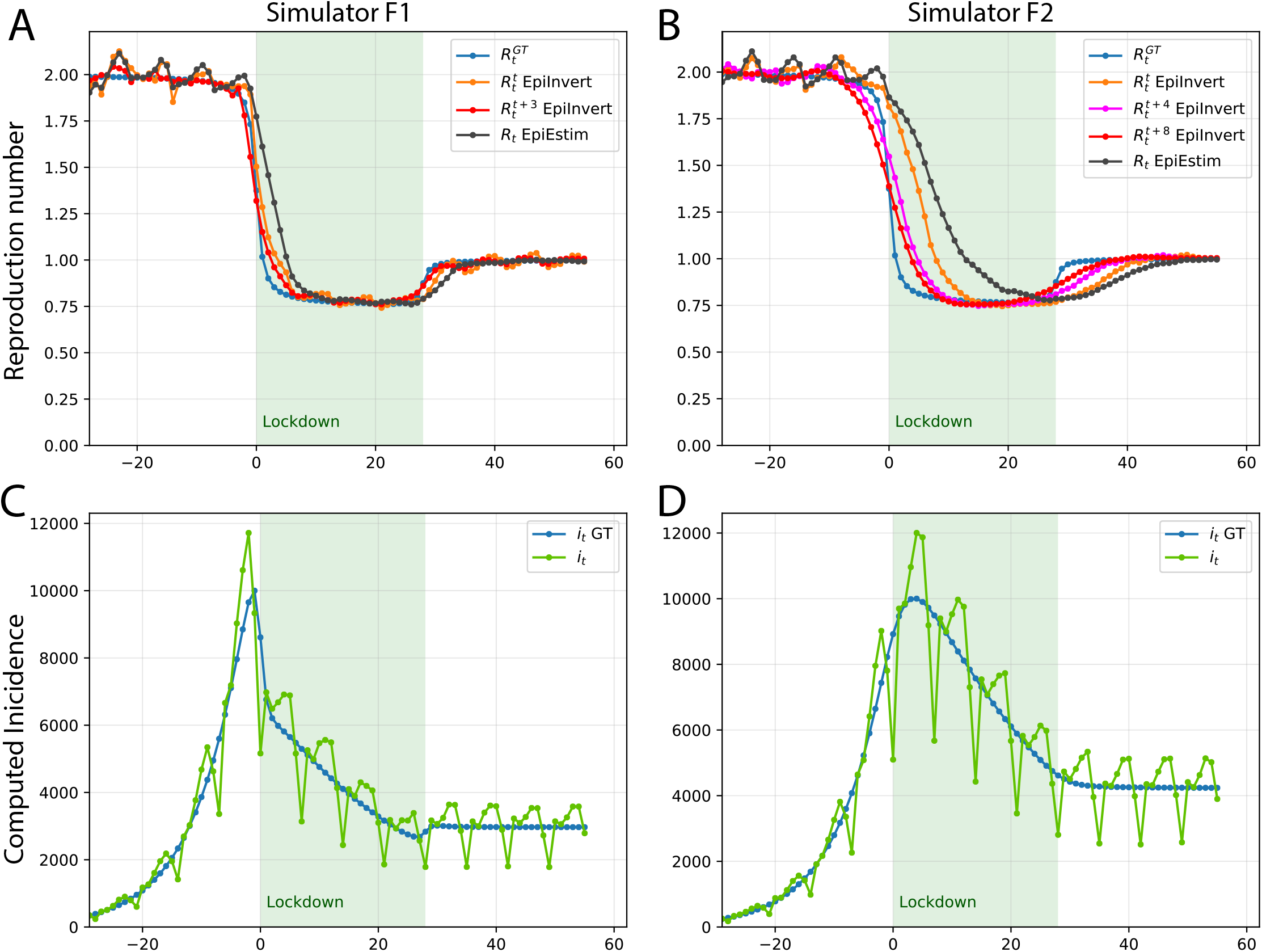
Comparison on a lockdown scenario of the results of the *R*_*t*_ estimation methods. The 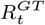 ground truth parameters are *R*_0_ = 2 (reproduction number before the lockdown), *R*_*i*_ = 0.75 (reproduction number after the lockdown), *t*′ = 28 (lockdown duration), *s* = 0.5 (slope of the transition between *R*_0_ and *R*_*i*_), *i*_*max*_ = 10000 (maximum of the incidence curve), and a weekly periodic bias borrowed from the USA. The simulations and inversions were performed in A and C for *F F*_1_ and in B and D for *F* ≡ *F*_2_. Note that the same 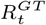 scenario (blue curve in A and B) leads to very different incidence curves (in green) in C an D. Hence, the results of the *F* 1 and *F* 2 inversions in A and B cannot be compared. In A and B, 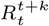 denotes the EpiInvert estimate of *R*_*t*_ obtained *k* days later.

**Fig. 10.**
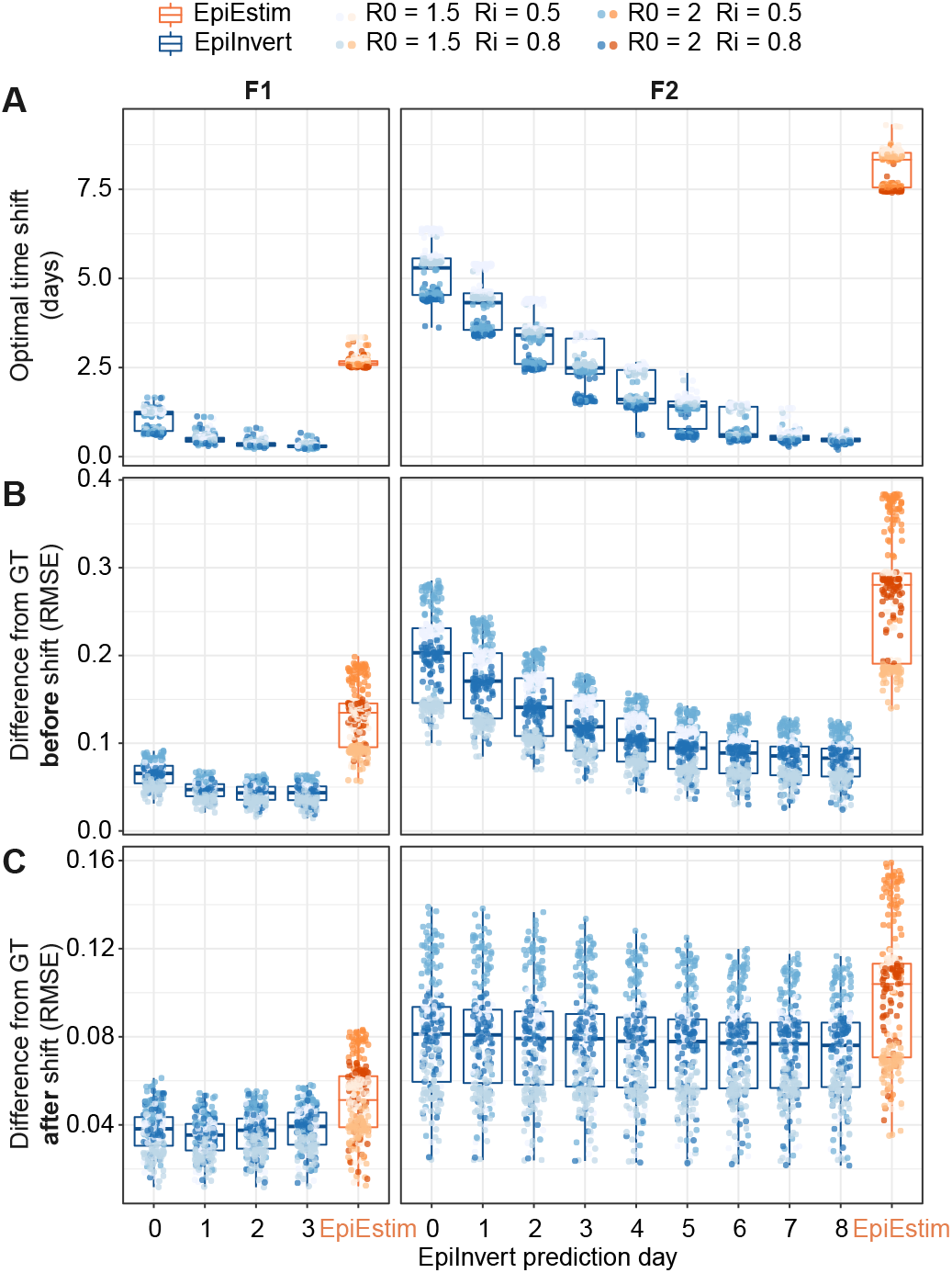
Distributions of the optimal time shift (A), the RMSE before the time shift (B) and the RMSE after the time shift (C) between the ground truth 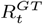 and its approximation obtained by EpiEstim and by EpiInvert, as a function of the number of days *k* after *t* used in the prediction. Note that columns F1 and F2 cannot be compared. Indeed, as illustrated in Figure 9, the incidence simulations for a same ground truth 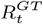 are quite different. The result of EpinInvert, which evolves with time, converges near-perfectly to the ground truth after 8 days (resp. 3 days) for F2 and F1 respectively. The EpiEstim result is static and stays 30 to 40% away from the ground truth. As argued in section 1, this large relative error in the F2 model can be compensated by shifting back the estimate by 8.5 days.

In Table 1, we show the distributions of the optimal time delay between 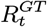 and its various estimates by minimizing the RMSE between both curves. For the F1 model, the EpiEstim estimate shifted back by 2.65 days has an error of 0.053. 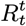, computed on the same day by EpinInvert, has an error of 0.044 with a delay of just 0.84 days. In short, EpiInvert gets a better estimate 2 days in advance with respect to EpiEstim. A similar conclusion arises for the *F*_2_ model. EpiEstim, when shifted back by 8.42 days, has an error of 0.108. Waiting for 8 days and shifting back by 0.44 days the result of 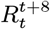 yields an inferior error, 0.075. But a result almost as good (0.078) is obtained by taking the result of 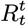 and shifting it back by 5.41 days. There is no particular gain in waiting longer for better estimates of EpinInvert: the estimate does not improve with time and is stack at 0.075. In summary, this result (based on simulations) leads to the following recommendations:

**Table 1.**
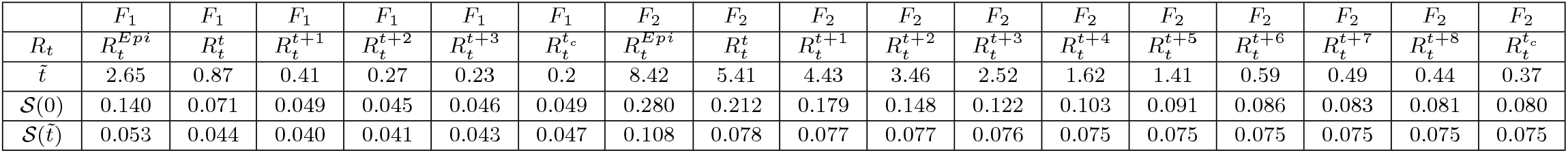
Statistical results for the different estimation results of *R*_*t*_ on 4800 simulations for the F1 and F2 models. First row: the renewal equation model used. Second Row: the *R*_*t*_ estimate. Third row: the optimal shift 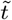 minimizing the RMSE between 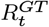 and the *R*_*t*_ estimate. Fourth row: the median of *𝒮* (0), the RMSE without temporal shift. Fifth row: the RMSE 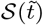 after applying the optimal temporal shift.

a. The EpiEstim estimate at time *t, R*_*t*_ must be shifted back by 8.42 days;
b. The EpinInvert synchronous estimate 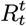 made at time *t* must be shifted back by 5.41 days. It is more precise than the EpiEstim estimate (an 0.078 error against 0.108) and it is obtained three days earlier (a 5.41 days delay against 8.42);
c. Nevertheless, as we have seen in Fig. 6, the EpiInvert ex post estimate 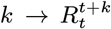 stabilizes after 5 days to a value which is very close to the ground truth, without the need for shifting back its value.

In the above estimates, replacing 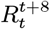 by 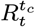 does not change this conclusion. The difference between these estimates is negligible. Indeed, 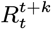 no longer varies for *k* ≥ 8.

## 5. CONCLUSION

The reproduction number *R*_*t*_ can be estimated by solving a renewal equation linking *R*_*t*_, the case incidence curve *i*_*t*_, and the serial interval Φ_*s*_. We considered two formulations of the renewal equation. The first one (*F* ≡ *F*_1_) estimates the instantaneous reproduction number. The second one (*F* ≡ *F*_2_) estimates the case reproductive number. Resolving these equations is challenging, because the daily incidence data *i*_*t*_ recorded by health administrations is noisy and shows a strong quasi-periodic behavior. In order to get an estimate of *R*_*t*_ we introduced a classic regularity constraint on *R*_*t*_ and we corrected the weekly periodic bias observed in the incidence curve *i*_*t*_ by a simple variational formulation. Our proposed variational model, EpiInvert, also computes empirical confidence intervals. In contrast to former methods, EpiInvert can use serial intervals with distributions containing negative days (as it is the case for COVID-19). Thus, it avoids an artificial truncation of the serial interval, and it provides an estimate that improves with time. Nevertheless, as shown on simulations and on real incidence data, EpiInvert shows excellent agreement with EpiEstim. Its main improvement is the reduction of the time shift between the estimation and the actual value of *R*_*t*_. If we compare EpiEstim with the EpiInvert estimate 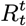 (the estimation of *R*_*t*_ that uses the incidence values up to the time *t*), EpiInvert provides an estimate of *R*_*t*_ about 2 days in advance for the instantaneous reproduction number, and about 3 days in advance for the case reproductive number. This means that for both models, EpiInvert can anticipate by several days an estimate of *R*_*t*_. This estimate is more precise than the EpiEstim estimate. In addition, we proved on a simulator that the EpiInvert estimate 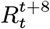, obtained 8 days later than the current date, *t*, is very close to the ground truth. Comparing it with the EpiEstim estimate confirms that the time delay of EpiEstim is about 3 days for the instantaneous reproduction number (*F* = *F*_1_), and more than 8 days for the case reproductive number (*F* = *F*_2_). Finally, comparing the EpiEstim and the EpiInvert estimated curves of *R*_*t*_ on real data confirms these 3 and 8 days delays between both estimates. These facts are extremely relevant, given that the control of social distancing policies requires a timely estimate of *R*_*t*_.

## Data Availability

We use the COVID-19 registered daily infected from https://ourworldindata.org

## Supporting Information

In this section we describe and analyze the EpiEstim method and its parameters (Section A). In Section B the Wallinga-Teunis method. Section C presents implementation details of EpiInvert. Section D shows some technical details on our experiments on simulated data. Section D makes a case study of the USA, France, Japan, Peru and South Africa.

### A. The EpiEstim method

One of the most widely used methods to estimate the instantaneous reproduction number is the EpiEstim method proposed by Cori et al. (5). In what follows, we will denote by 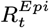 the EpiEstim estimation. The authors show that if *i*_*t*_ follows a Poisson distribution with expectation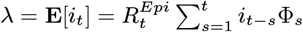 and 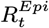 is assumed to follow a gamma prior distribution Γ(*a, b*), then the following analytical expression can be obtained for the posterior distribution of 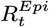:

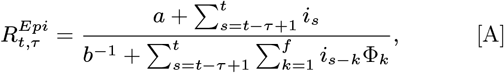

where 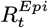 is assumed to be locally constant in a time window of size *τ* ending at time *t*. However, *i*_*t*_ does not follow a Poisson distribution as its local variance in most states much higher than its mean, being dominated by the weekend effect. In this method, implemented in the EpiEstim R package, a regularization of the estimation is introduced by assuming that 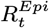 is constant in a time window of size *τ* ending at time *t*. We found that the parameters *a* and *b* of the prior Gamma distribution Γ(*a, b*), have very little influence on the current estimation of 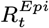. Cori et al. in (5) proposed to use *a* = 1 and *b* = 5. Taking into account the magnitude of the current number of daily cases in countries affected by Covid-19, the contribution of *a* and *b* to the expression [A] can be neglected. As shown in (15), assuming that the mean *ab* of the prior Gamma distribution Γ(*a, b*) satisfies

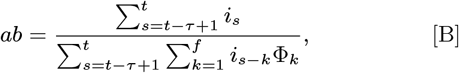

equation [A] becomes

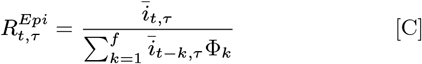

which corresponds to the usual 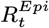 estimate obtained directly from equation [2] applied to 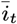, where 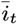 is the average of *i*_*t*_ in the interval [*t* −*τ, t*]. Therefore, if we remove the parameters *a* and *b* from the estimation of 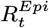, the main difference between the EpiEstim estimation and the one proposed here for *F* ≡ *F*_1_ is that in EpiEstim, a serial interval with non-positive values is not allowed and that the regularity is forced by a backward seven day average of the incidence curve. This is replaced by a regularity term in the proposed variational formulation. Notice that due to the backward averaging of the incidence curve, we can expect a time shift between both estimations.

### B. The Wallinga and Teunis computation of *R*_*t*_

The Wallinga-Teunis (4) method is also implemented in the EpiEstim package and widely considered as a reliable method to compute the case reproduction number, 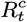, retrospectively (11). Its formulas to estimate 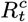 at time *t* require the knowledge of *i*_*t*_ for *t* = 0, *…, t* + *f*. Starting from the original definitions of the authors, we give a mathematical proof that their method is actually computing *R*_*t*_ by the *F*_1_ form of the renewal equation. The method is based on the following estimation of the “relative likelihood, *p*_*k,l*_, that a case *k* has been infected by case *l*”,

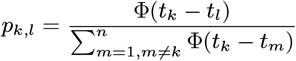

where *n* represents the reported cases and *t*_*k*_ is the time of infection for the case *k*. Wallinga and Teunis define the *case reproduction number* by

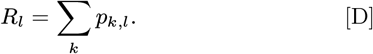

Since *R*_*l*_ only depends on the time of infection *t*_*l*_, it is actually an estimation of the reproduction number at time *t* = *t*_*l*_, so the Wallinga and Teunis estimate, 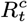, of the reproduction number can be expressed as:

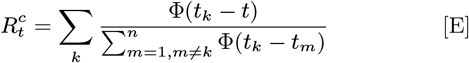

It remains to establish a relation of 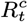 with the instantaneous reproduction number 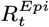 obtained by the renewal equation with *F* ≡ *F*_1_,

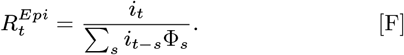

Grouping in the sum in [E] the cases *k* such that 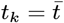 and taking into account that there are 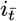 such cases, 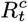 can be rewritten as

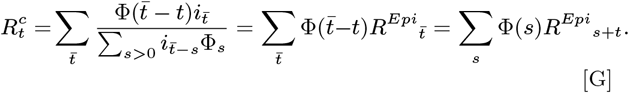

We can therefore interpret 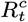 as the forward convolution of the initial estimate 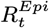 with the kernel given by Φ_*s*_. This relation between the instantaneous and case reproduction numbers has also been proven in (9) (equation (10)). On the other hand, as explained above, the EpiEstim estimate 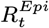 can be interpreted (if we neglect the parameters *a* and *b* of the Gamma distribution) as the application of Equation [F] to the incidence curve filtered by sliding average on [*t* − *τ* + 1, *t*]. In conclusion the Cori et al. and the Wallinga and Teunis methods use the renewal equation *F* ≡ *F*_1_. Note, however, that the Wallinga and Teunis method computes the reproduction number only retrospectively. Indeed, the computation of 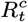 requires the values of 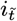 for any 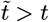 such that 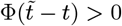. This fact was observed in Cori et al.: (in the WT approach), “estimates are right censored, because the estimate of *R* at time *t* requires incidence data from times later than *t*.”

### C. Implementation details of EpiInvert

#### Boundary condition for [t > t_c_]

The proposed inversion model provides an estimation of *R*_*t*_ up to the the date,*t*_*c*_, of the last available incidence value. An obvious objection is that if *f*_0_ *<* 0, the functional [9] involves a few future values of *R*_*t*_ and *i*_*t*_ for *t*_*c*_ ≤*t* ≤*t*_*c*_ −*f*_0_. These values are unknown at present time *t*_*c*_. We use a basic linear regression to extrapolate the values of *i*_*t*_ beyond *t*_*c*_. To compute the regression line (*i* = *m*_7_ · *t* + *n*_7_) we use the last seven values of *i*_*t*_. In summary, the extension of *i*_*t*_ beyond the observed interval [0, *t*_*c*_] is defined by

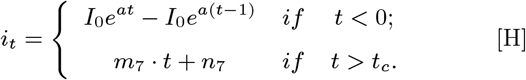

The above defined boundary conditions has a very minor influence in the final estimation of *R*_*t*_ in the last days when minimizing [9]. Indeed, the extension of *i*_*t*_ for *t <* 0 is only relevant at the beginning of the epidemic spread. On the other hand, the extension of *i*_*t*_ for *t > t*_*c*_ is only required when the serial interval has negative values. For instance, to evaluate the renewal equation in the energy at the current time *t*_*c*_ using this approach for *F* ≡ *F*_2_ we use the expression

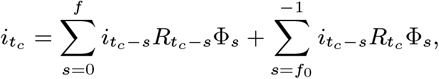

and the extension of *i*_*t*_ for *t > t*_*c*_ is only used in the last term of the above expression where the values of Φ_*s*_ are usually very small. Hence the influence of this extension procedure for *i*_*t*_ is also almost negligible. To confirm this claim, we compared, using the shifted log-normal approximation of the serial interval proposed by Ma et al., the estimate of 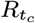 using the extrapolation based on a linear regression of the last 7 days, with the basic extrapolation given by 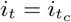 for *t > t*_*c*_. Computing the absolute value of the difference of both estimates for 81 countries we obtain that the quartiles of such distribution of values are *Q*_0_ = 6.6 ·10^−6^, *Q*_1_ = 1.3 ·10^−4^, *Q*_2_ = 3.1 ·10^−4^, *Q*_3_ = 5.7 ·10^−4^ and *Q*_4_ = 4.9 ·10^−3^. We conclude that extrapolation of *i*_*t*_ beyond *t*_*c*_ is a valid strategy to estimate *R*_*t*_ up to *t* = *t*_*c*_.

#### Pre-processing the incidence curve

Some countries do not provide data on holidays or weekends and only provide the cumulative total of cases on the next working day. To avoid the strong discontinuity in the data sequence produced by the lack of data, we automatically divide the case numbers of the first non-missing day, between the number of days affected. We do not allow negative numbers in the incidence curve. By default, we replace by zero any negative value of the incidence curve.

#### Alternate minimization of the energy [9]

To minimize the energy [9], we use an alternate minimization algorithm with respect to *R*_*t*_ and **q**. Indeed, if **q** is fixed, then the optimization of the energy [9] with respect to *R*_*t*_ leads to a linear system of equations that is easily solved. In what follows, we will denote by *R*(*t, i*, **q**) the result of this minimization. On the other hand, when *R*_*t*_ is fixed, the minimization of [9] with respect to **q** also leads to a linear system of equations. The constraint [10] is expressed as an additional linear equation,

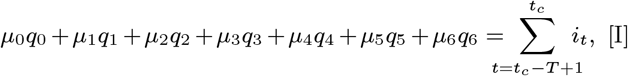

where 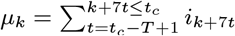. This linear constraint is easily included in the minimization procedure using, for instance, Lagrange multipliers. So **q** is computed as the unique solution of the associated linear system. In what follows we will denote by **q**(*R*) the result of this minimization. Let us denote by 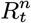 and **q**^*n*^ the estimation of *R*_*t*_ and **q** in the iteration *n* of the alternate minimization algorithm. We also denote by 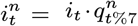 the filtered incidence curve at iteration *n*. We initialize *n* = 0, *i*^0^ ≡*i*, **q**^0^ ≡1 and we compute 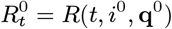 as the minimizer of the energy [9] with respect to *R*_*t*_ for **q** ≡**q**^0^.

The whole method is summarized in Algorithm 1, where the maximum number of iterations is fixed to *MaxIter* = 100.

##### Algorithm 1

Estimation of 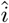, *R*, **q** from *i* and Φ.

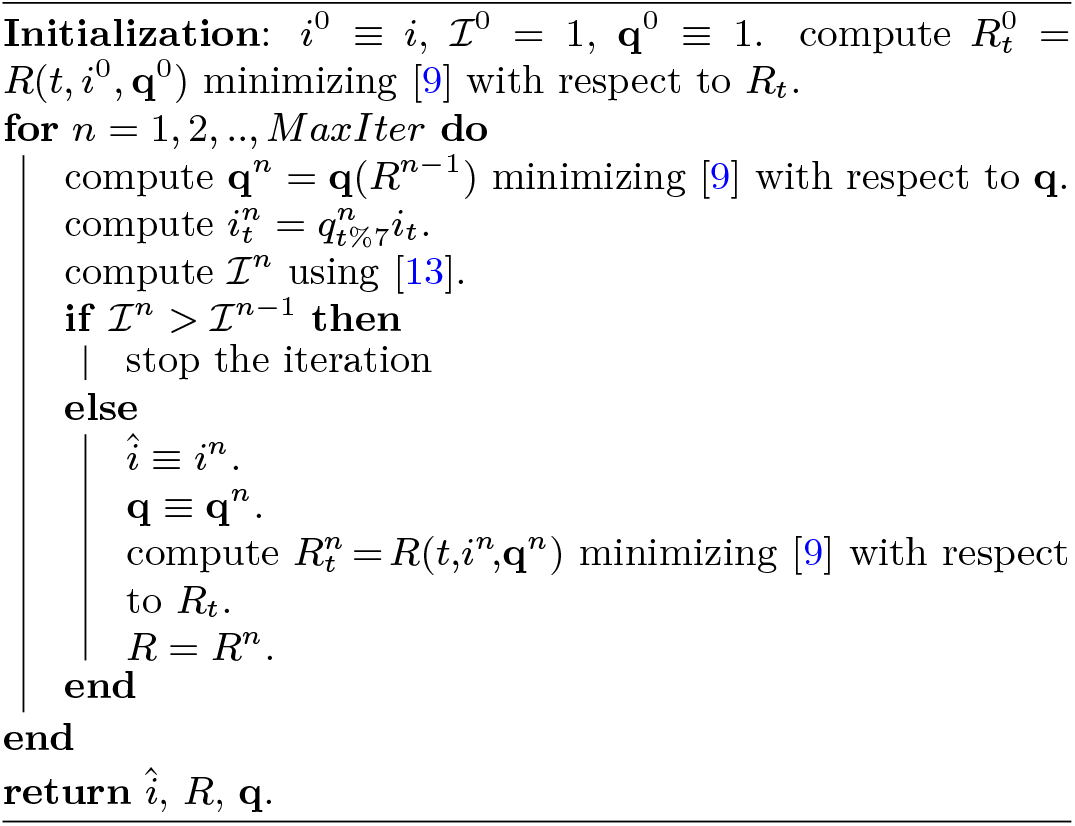

#### *Initial boundary condition, for t* = 0

The evaluation of *F*_2_(*i, R*, Φ, *t*) requires values of *R*_*t*_ and *i*_*t*_ beyond the interval [0, *t*_*c*_]. Given the boundary conditions established, we assume that *R*_*t*_ = *R*0 for *t <* 0 and 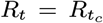 for *t > t*_*c*_. Concerning *i*_*t*_, for *t <* 0 we will assume, as usual, that at the beginning of the epidemic spread the virus is in free circulation and the cumulative number of infected detected 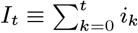 follows an exponential growth for *t <* 0, that is *I*_*t*_ = *I*_0_*e*^*at*^, where *a* represents the initial exponential growth rate of *I*_*t*_ at the beginning of the infection spread. We now naturally estimate *a* by

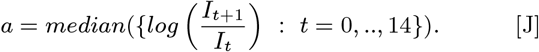

If we assume that *I*_*t*_ = *I*_0_*e*^*at*^ follows initially an exponential growth and that *R*_*t*_ = *R*0 is initially constant, then we can compute *R*0 from the exponential growth *a* and the renewal equation taking into account that

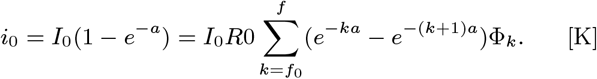

Hence, we can compute an approximation of *R*0 as

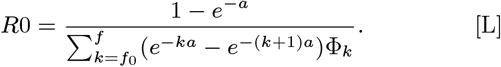

This estimation of *R*0 is a discrete version of the formula given in (9) where the incidence curve is assumed to follow an exponential growth. Note that this estimation strongly depends on the serial interval used. For instance, if we assume that *a* = 0.250737 (the exponential growth rate obtained in (25) when the coronavirus is in free circulation), we obtain that *R*0 = 2.700635 for the Nishiura et al. serial interval, *R*0 = 3.084528 for the Ma et al. serial interval and *R*0 = 1.839132 for the Du et al. serial interval.

### D. Experiments using simulated data

We describe here in more detail a simulator *R*_*t*_ →*i*_*t*_. The simulator starts from a realistic scenario on the evolution of *R*_*t*_ depending on parameters fixed by the user. Then, using a choice of serial intervals and a realistic weekly bias borrowed from real examples, the simulator samples the incidence curve *i*_*t*_ as the realization of Poisson variable. The simulated “ground truths” for *R*_*t*_ will be denoted by 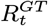. They are similar to those proposed in Gostic et al. (11). 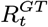 goes from a user selected initial value *R*_0_ *>* 1 to an intermediate value *R*_*i*_ *<* 1, and finally goes back to 1. This hypothesis for an evolution corresponds to a typical lockdown scenario where initially the number of cases grows exponentially, then a lockdown is implemented during a time that we denote by *t′*, and finally social-distancing measures relax but try to stabilize *R*_*t*_ around 1. The parameters defining the simulated ground truth 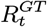 therefore are *R*_0_, *R*_*i*_, *t′* and *s*, which determines the slope of the transitions between *R*_0_ and *R*_*i*_ and between *R*_*i*_ and 1. the larger *s*, the sharper the transitions. To define 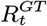 we use the following function:

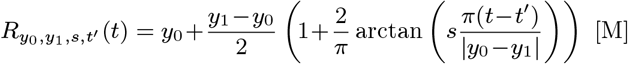

where *y*_0_, *y*_1_, *s* and *t′* are the function parameters. This function satisfies: 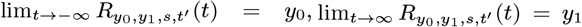. The maximum of the absolute value of its derivative is equal to *s* and is attained at *t* = *t′*. Next, we define 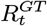 by

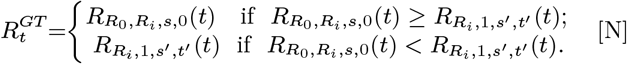

To reduce the number of parameters we assume that *s′* = *s/*5, reflecting the fact that the relaxation of social distancing measures is more progressive than a lockdown.

The ground truth of the incidence curve, that we denote by 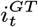, is computed from the renewal equation using 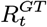 as reproduction number and a user selected serial interval among three proposed (Du, Ma, Nishiura). We take an initial value for 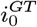 and iteratively compute 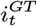 from 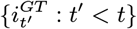 using the renewal equation and the boundary conditions explained above. Then, we improve the accuracy of the estimation of 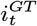 by applying a Newton method until convergence. Indeed, given 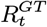, the renewal equation is a fixed point equation in *i*_*t*_. Since the ground truth of the incidence curve is defined up to the multiplication by a constant factor, rather than fixing the initial number of cases, we add a more intuitive parameter *i*_*max*_ which allows the user to fix the maximum value of the incidence curve in the whole period. This value impacts the noise inherent to a Poisson process: the smaller *i*_*max*_, the larger the stochastic oscillation of *i*_*t*_. We then simulate the observed incidence curve *i*_*t*_ assuming that 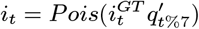, that is, *i*_*t*_ follows a Poisson distribution of mean 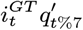 where 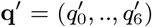 is the vector with the weekly bias correction factors. The weekly bias proposes several real bias correction factors options **q** = (*q*_0_, .., *q*_6_), borrowed from the EpinInvert analysis of the incidence curve of 19 countries. To obtain **q′** from **q**, we simply invert the weekly bias correction coefficients by setting 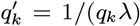, where *λ* is a normalization factor preserving the cumulative number of cases in the period of analysis. More precisely, *λ* is derived from the equation

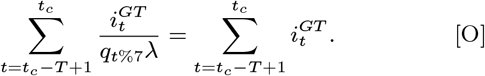

**Fig. S1.**
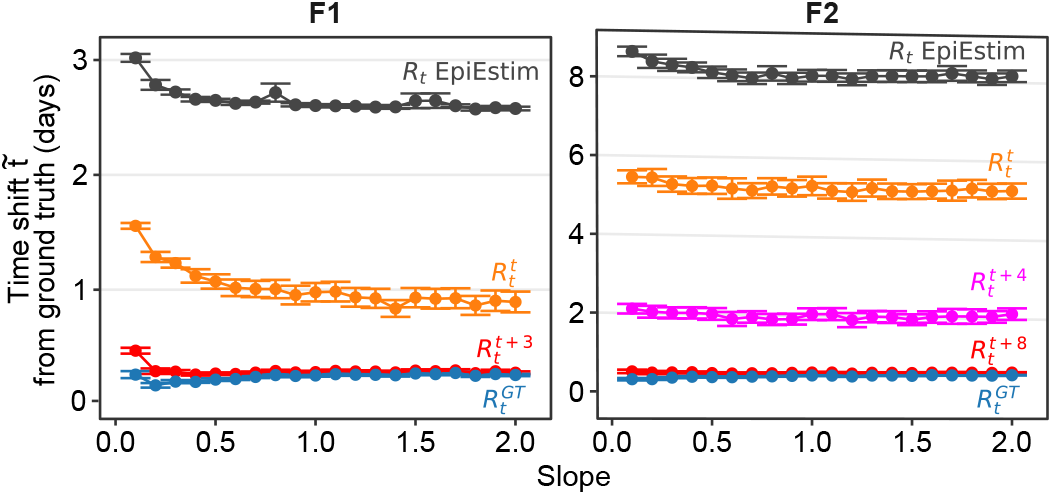
Time delay between the reproduction number ground truth 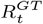 and its various estimates when computing the time shift that minimizes the RMSE between both curves. The horizontal axis is the slope of *R*_*t*_ at lockdown time (*t* = 0). Using the simulator, these estimates confirm the 8.5 days delay of the EpiEstim estimate with respect to the ground truth for F2, and a 3 days delay for the F1 form of the equation. The EpiInvert delay is also important on the first evaluation day, but decreases as days pass by.

From a sample of the stochastic simulation of *i*_*t*_, the demo finally computes *R*_*t*_ using EpiInvert and EpiEstim.

In Fig. S1 we show, as a function of the slope in the lockdown transition, the distributions of the optimal time delay between 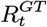 and its various estimates. The optimal time shift between an *R*_*t*_ curve and 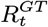 is the one that minimizes their RMSE. We observe that the time shift is slightly larger when the slope of the transition is small.

### E. Case studies: USA, France, Japan, Peru and South Africa

The country data about the registered daily infected are taken from https://ourworldindata.org. In the particular cases of France, Spain and Germany we use the official data reported by the countries. We shall use the incidence data up to Friday, July 23, 2021 (so the last weekly bias correction factor *q*_6_ corresponds to a Friday). For the US states, the data are obtained from the New York Times report ^§^.

Table S2 contains a summary of the values computed for each experiment. To compute the EpiEstim estimation 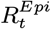, we used *τ* = 7, that is, we assumed that *R*_*t*_ is constant in [*t* −7, *t*]. As proposed by Cori et al. in (5) we used *a* = 1 and *b* = 5 for the parameters of the Γ(*a, b*) prior distribution for *R*_*t*_. Yet, as explained above, these values could be neglected in the EpiEstim estimation, given the magnitude of the incidence data in these regions. The values of the bias correction coefficients *q*_*k*_ obtained for *F* ≡*F*_1_ and *F* ≡ *F*_2_ are quite similar. So it seems that the choice of the renewal equation has no significant influence on the estimation of the bias correction coefficients.

In Fig. S2 we show the charts obtained for the USA with *F* ≡*F*_1_ and *F* ≡ *F*_2_. The USA shows a clear weekly periodic bias. The correction of this bias works quite well, as the RMSE reduction reaches ℐ = 0.409 for *F* ≡ *F*_1_ and ℐ = 0.381 for *F* ≡ *F*_2_. The oscillation of the incidence curve is strongly reduced, passing from 𝒱 (*i*) = 0.542 to 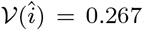. The agreement with EpiEstim is also excellent as 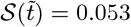 for *F* ≡ *F*_1_ and 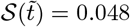 for *F* ≡ *F*_2_. The daily bias correction factors are similar for *F* ≡ *F*_1_ and *F* ≡ *F*_2_. On Sundays the number of cases is strongly underestimated (*q*_1_ = 3.205 for *F* ≡ *F*_2_) and overestimated on Fridays (*q*_6_ = 0.569 for *F*≡*F*_2_).

**Table S2.**
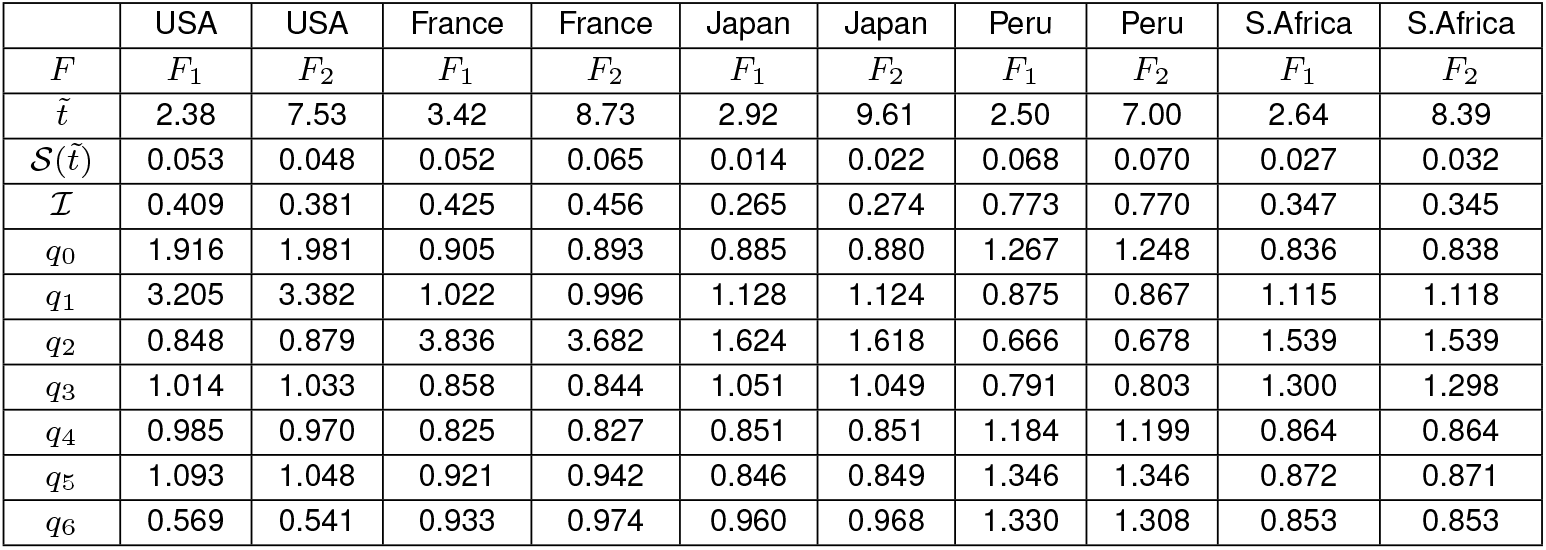
Numerical results obtained by EpiInvert for the USA, France, Japan, Peru and South Africa using the data up to July 23, 2021 and the renewal equations *F* = *F*_1_ and *F* = *F*_2_.

In Fig. S3 we show the charts obtained for France with *F*≡*F*_1_ and *F*≡*F*_2_. France also displays a clear weekly periodic bias: on Mondays the number of cases is strongly underestimated (*q*_2_ = 3.682 for *F*≡*F*_2_). The correction of the periodic bias works well, as ℐ = 0.425 for *F*≡*F*_1_ and ℐ = 0.456 for *F*≡*F*_2_. The oscillation of the incidence curve is therefore reduced, passing from 𝒱 (*i*) = 0.346 to 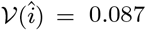. The agreement with the EpiEstim method is good, with 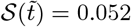 for *F*≡*F*_1_ and 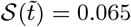 for *F*≡*F*_2_.

In Fig. S4 we show the charts obtained for Japan with *F*≡*F*_1_ and *F*≡*F*_2_. In Japan, the weekly bias correction works very well and yields ℐ = 0.265 for *F*≡*F*_1_ and ℐ = 0.274 for *F*≡*F*_2_. The oscillation of the incidence curve reduces from 𝒱 (*i*) = 0.189 to 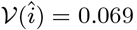. The agreement with the EpiEstim method is good, with 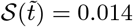 for *F*≡*F*_1_ and 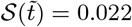 for *F*≡*F*_2_. Observe how the incidence curve is underestimated on Mondays (*q*_2_ = 1.618).

In Fig. S5 we show the charts obtained for Peru with *F*≡*F*_1_ and *F*≡*F*_2_. Although in general countries present a clear weekly periodic pattern in the incidence curve this is not the case for Peru: we obtain ℐ = 0.773 for *F*≡*F*_1_ and ℐ = 0.770 for *F*≡*F*_2_. The oscillation of the incidence curve is not reduced, going from 𝒱(*i*) = 0.355 to 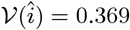. The agreement with EpiEstim is good with 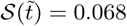 for *F*≡*F*_1_ and 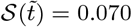 for *F*≡*F*_2_.

In Fig. S6 we show the charts obtained for South Africa with *F*≡*F*_1_ and *F*≡*F*_2_. The correction of the periodic bias works well, as ℐ = 0.347 for *F*≡*F*_1_ and ℐ = 0.345 for *F*≡*F*_2_. The oscillation of the incidence curve is reduced, passing from 𝒱 (*i*) = 0.191 to 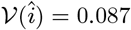. On Mondays the number of cases is underestimated (*q*_2_ = 1.539 for *F*≡*F*_2_). The agreement with the EpiEstim method is good, with 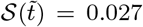 for *F*≡*F*_1_ and 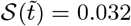 for *F*≡*F*_2_.

The optimal shift 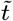 between *R*_*t*_ is 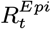 obtained for the different countries fits in the range obtained by a joint analysis of the 55 countries. Indeed, for 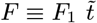 ranges from 2.38 to 3.42 and for 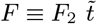 ranges from 7.00 to 9.61.

**Fig. S2.**
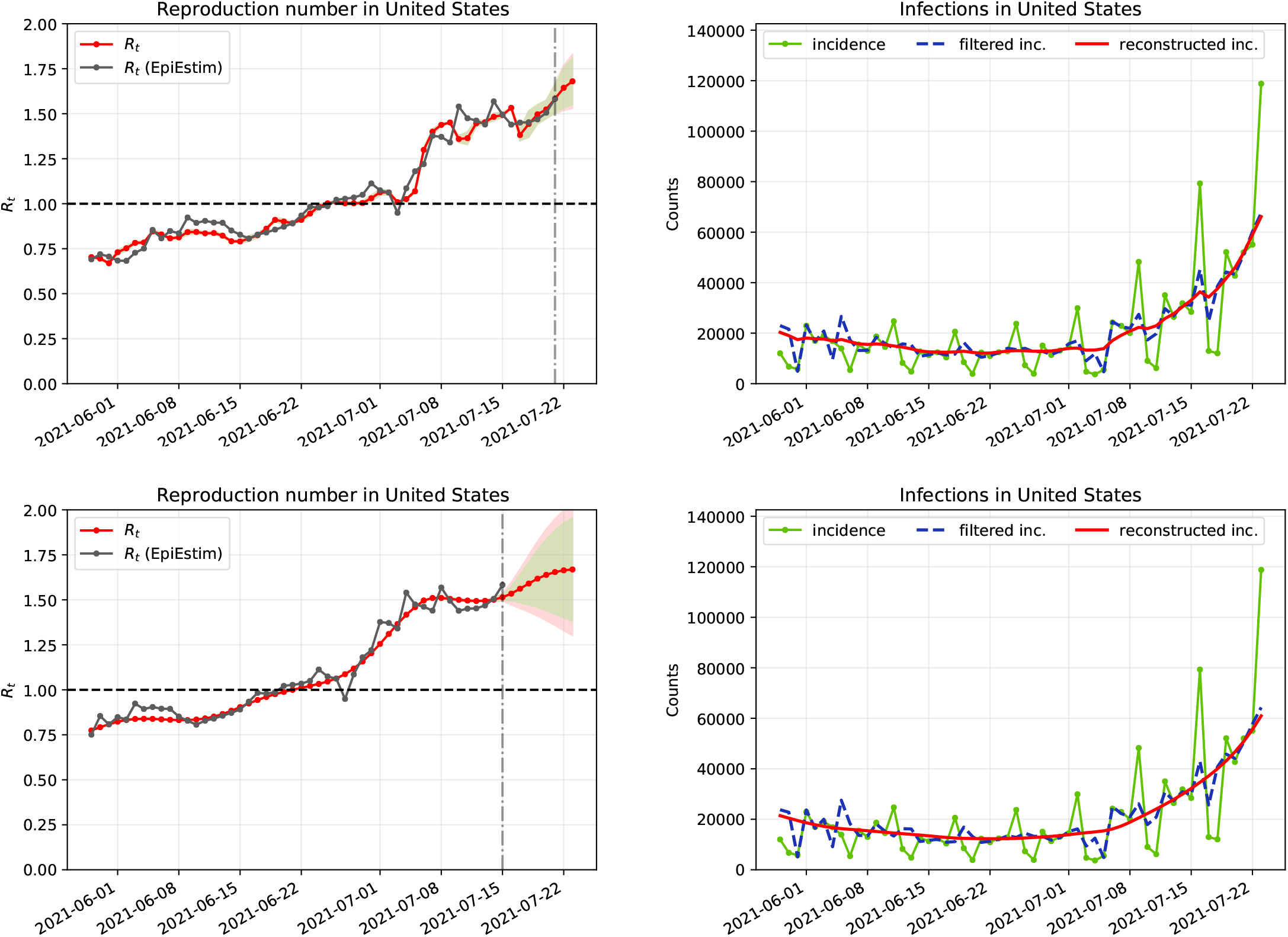
Results obtained for the USA up to July 23, 2021 using: (top) *F* ≡ *F*_1_ and (down) *F* ≡ *F*_2_.

**Fig. S3.**
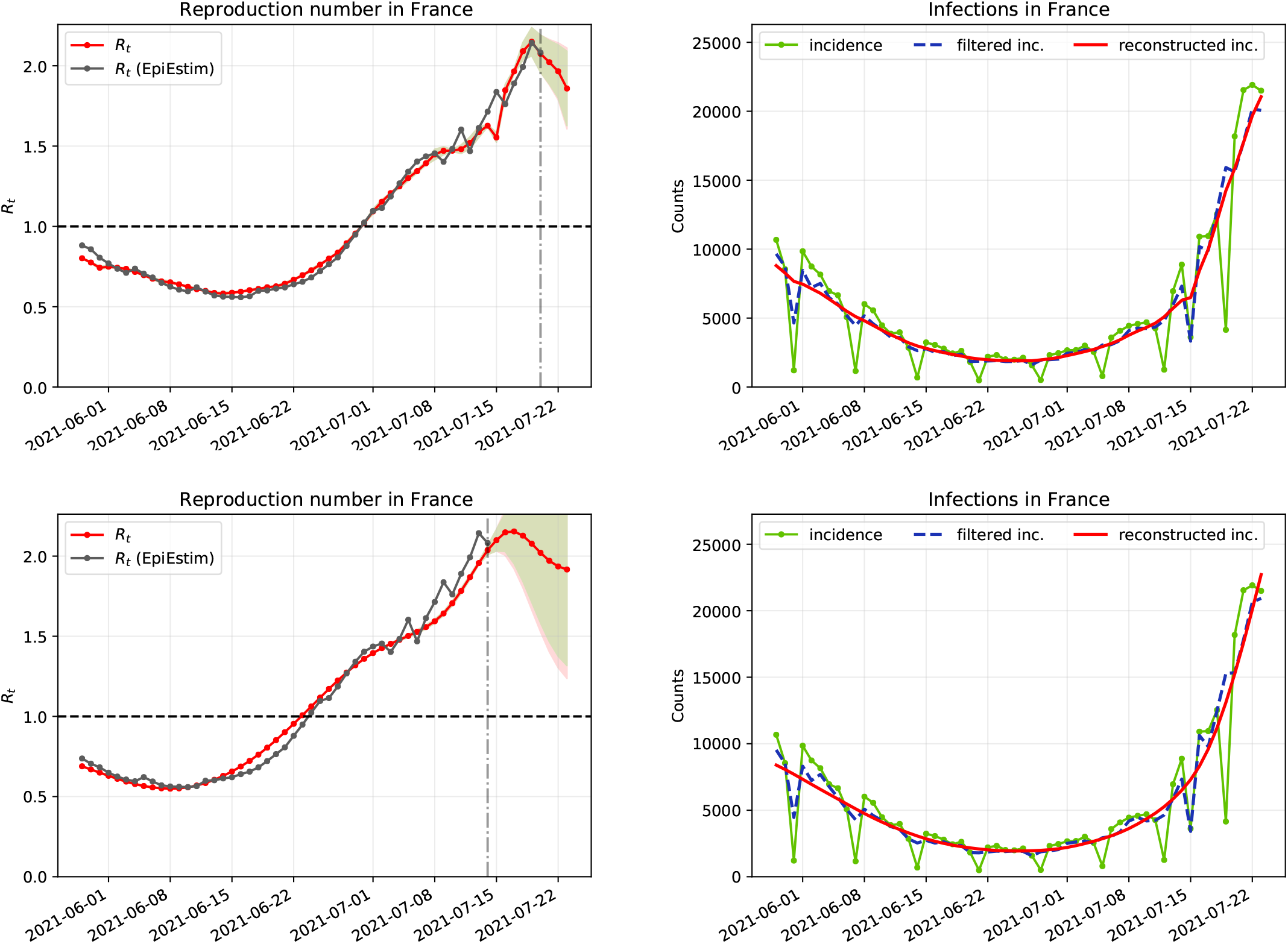
Results obtained for France up to July 23, 2021 using: (top) *F* ≡ *F*_1_ and (down) *F* ≡ *F*_2_.

**Fig. S4.**
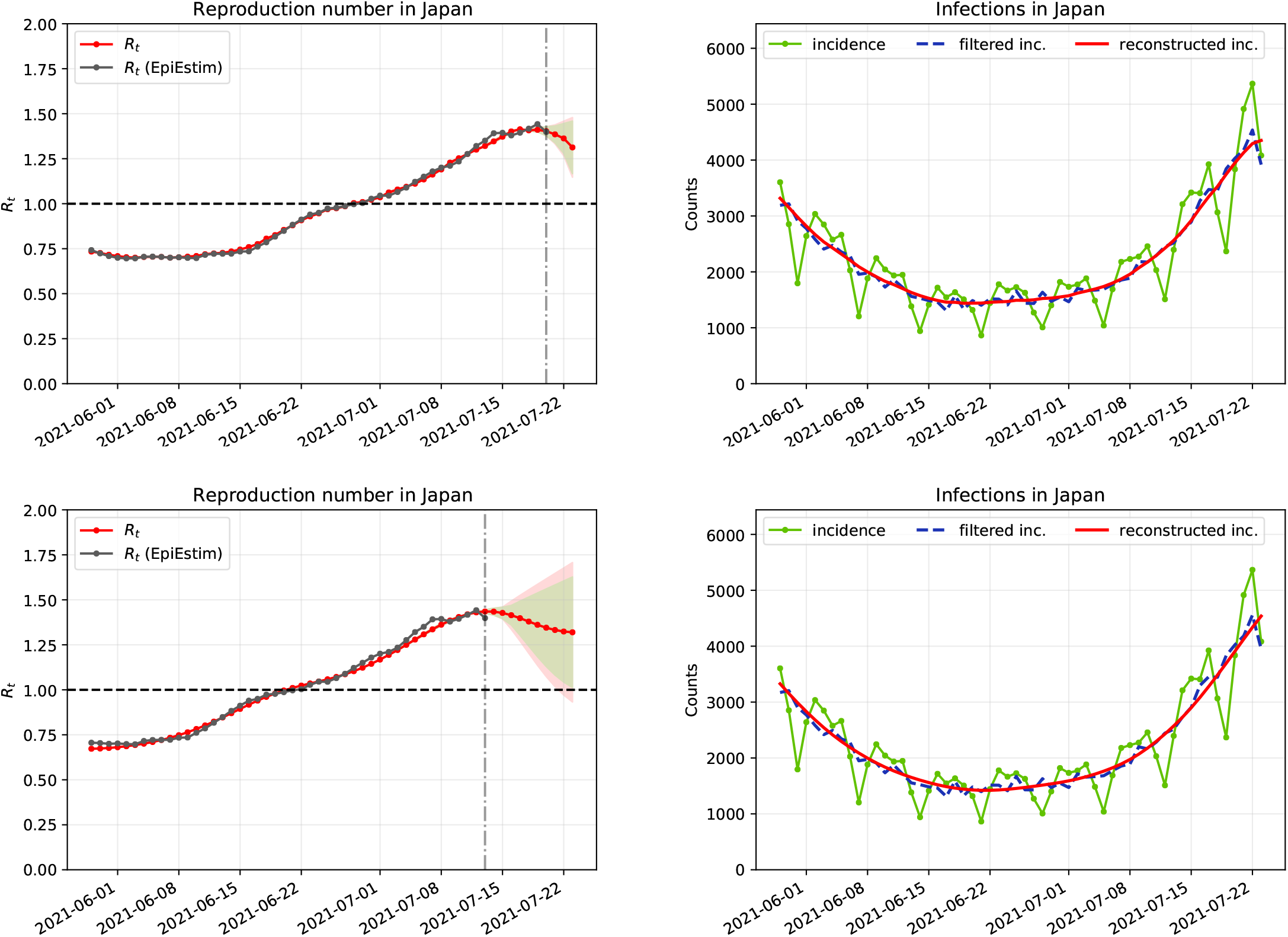
Results obtained for Japan up to July 23, 2021 using: (top) *F* ≡ *F*_1_ and (down) *F* ≡ *F*_2_.

**Fig. S5.**
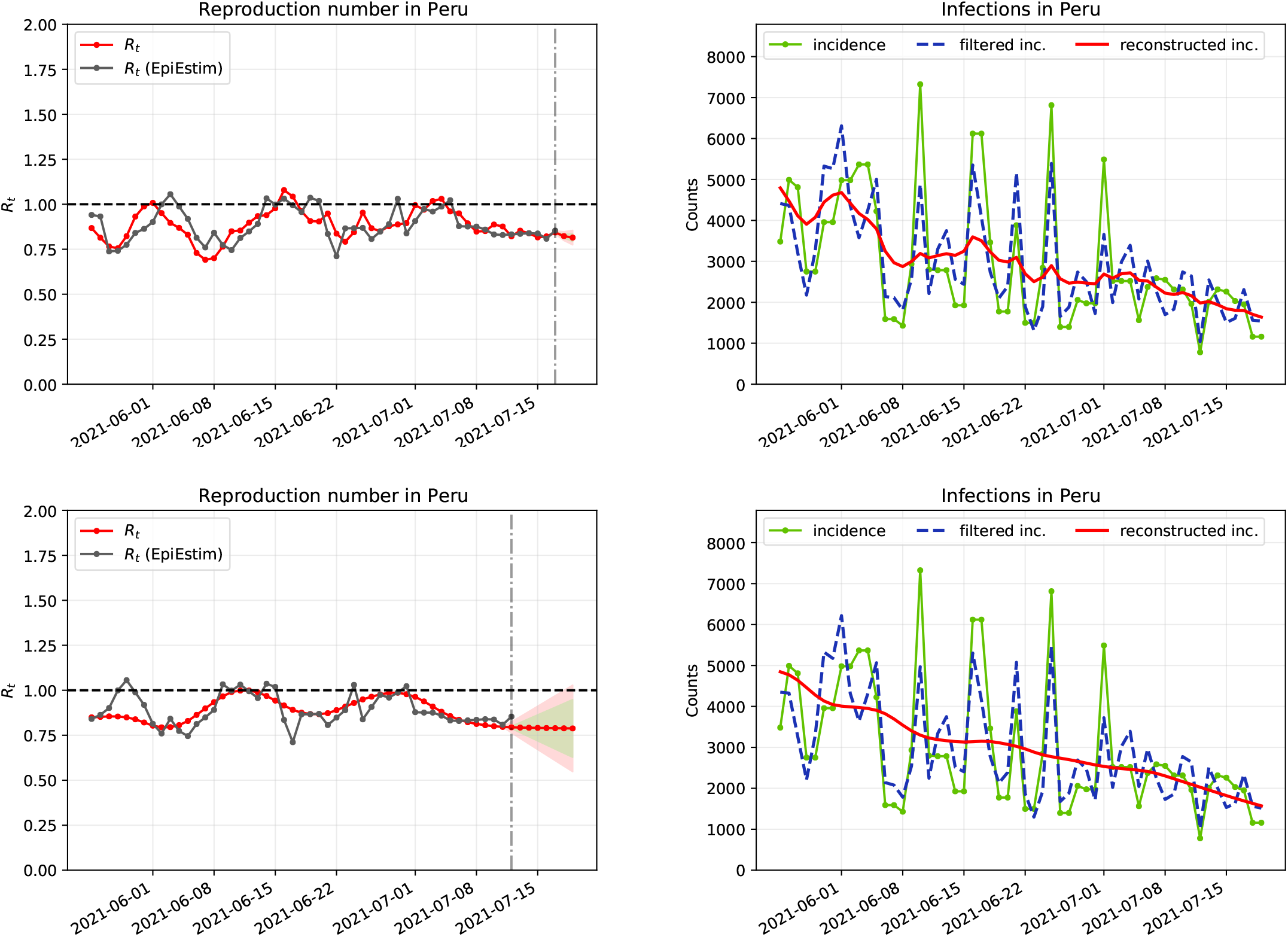
Results obtained for Peru up to July 23, 2021 using: (top) *F* ≡ *F*_1_ and (down) *F* ≡ *F*_2_.

**Fig. S6.**
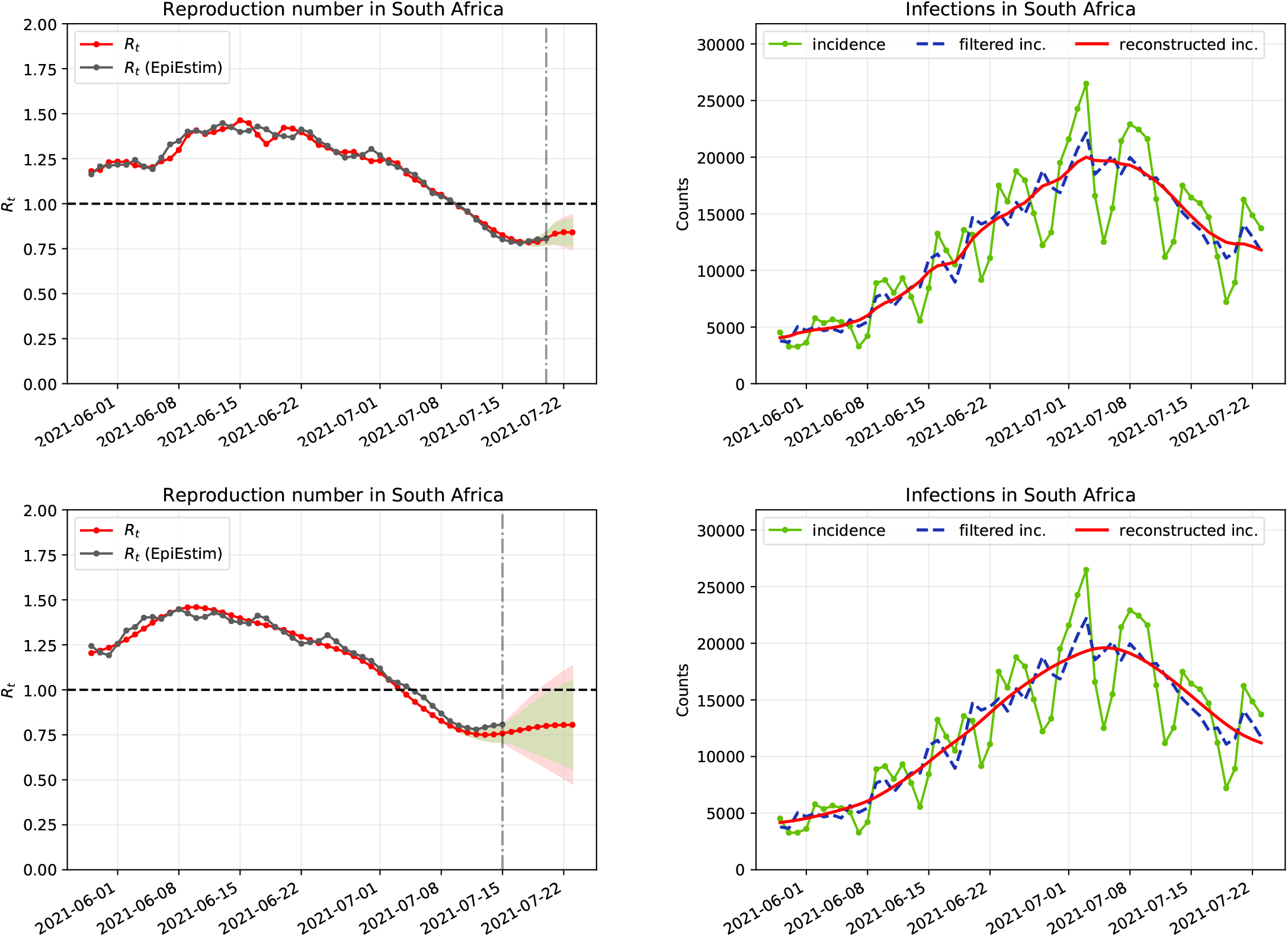
Results obtained for South Africa up to July 23, 2021 using: (top) *F* ≡ *F*_1_ and (down) *F* ≡ *F*_2_.

## Footnotes

* Cori et al. (5): “We assume that, once infected, individuals have an infectivity profile given by a probability distribution *w*_*s*_, dependent on time since infection of the case, *s*, but independent of calendar time, *t*. (…) *R*_*t*_ is the average number of secondary cases that each infected individual would infect if the conditions remained as they were at time *t*.”

† The lack of confidence in the computation of *R*_*t*_ is illustrated by the following fact: the official value of *R*_*t*_ is updated weekly and not daily by the official French online app Anticovid. This actually introduces an additional average 3.5 delay in the publication of this index!

‡ In the online interface (www.ipol.im/epiinvert) the users can, optionally, upload their own distribution for the serial interval.

§ https://raw.githubusercontent.com/nytimes/covid-19-data/master/us-states.csv

## References

1. P Rodpothong, P Auewarakul, Viral evolution and transmission effectiveness. World J. Virol. 1, 131 (2012).

2. X He, et al., Temporal dynamics in viral shedding and transmissibility of covid-19. Nat. medicine 26, 672–675 (2020).

3. P Ashcroft, et al., Covid-19 infectivity profile correction. arXiv preprint 2007.06602 (2020).

4. J Wallinga, P Teunis, Different epidemic curves for severe acute respiratory syndrome reveal similar impacts of control measures. Am. J. epidemiology 160, 509–516 (2004).

5. A Cori, NM Ferguson, C Fraser, S Cauchemez, A new framework and software to estimate time-varying reproduction numbers during epidemics. Am. journal epidemiology 178, 1505– 1512 (2013).

6. AJ Lotka, Relation between birth rates and death rates. Science 26, 21–22 (1907).

7. H Nishiura, Time variations in the transmissibility of pandemic influenza in Prussia, Germany, from 1918–19. Theor. Biol. Med. Model. 4, 20 (2007).

8. H Nishiura, G Chowell, The Effective Reproduction Number as a Prelude to Statistical Estimation of Time-Dependent Epidemic Trends, eds. G Chowell, JM Hyman, LMA Bettencourt, C Castillo-Chavez. (Springer Netherlands, Dordrecht), pp. 103–121 (2009).

9. C Fraser, Estimating individual and household reproduction numbers in an emerging epidemic. PLOS ONE 2, 1–12 (2007).

10. K Gostic, et al., Practical considerations for measuring the effective reproductive number, Rt. MedRxiv (2020).

11. KM Gostic, et al., Practical considerations for measuring the effective reproductive number, rt. PLoS computational biology 16, e1008409 (2020).

12. R Thompson, et al., Improved inference of time-varying reproduction numbers during infectious disease outbreaks. Epidemics 29, 100356 (2019).

13. QH Liu, et al., Measurability of the epidemic reproduction number in data-driven contact networks. Proc. Natl. Acad. Sci. 115, 12680–12685 (2018).

14. T Obadia, R Haneef, PY Boëlle, The r0 package: a toolbox to estimate reproduction numbers for epidemic outbreaks. BMC medical informatics decision making 12, 147 (2012).

15. TZ Boulmezaoud, L Alvarez, M Colom, JM Morel, A Daily Measure of the SARS-CoV-2 Effective Reproduction Number for all Countries. Image Processing On Line 10, 191–210 (2020) https://doi.org/10.5201/ipol.2020.304.

16. S Ma, et al., Epidemiological parameters of coronavirus disease 2019: a pooled analysis of publicly reported individual data of 1155 cases from seven countries. Medrxiv (2020).

17. J Demongeot, K Oshinubi, H Seligmann, F Thuderoz, Estimation of daily reproduction rates in covid-19 outbreak. medRxiv (2021).

18. AN Tikhonov, VY Arsenin, Solutions of ill-posed problems. New York 1, 30 (1977).

19. M Benning, M Burger, Modern regularization methods for inverse problems. Acta Numer. 27, 1–111 (2018).

20. J Asher, Forecasting ebola with a regression transmission model. Epidemics 22, 50–55 (2018) The RAPIDD Ebola Forecasting Challenge.

21. Z Du, et al., The serial interval of COVID-19 from publicly reported confirmed cases. medRxiv (2020).

22. H Nishiura, NM Linton, AR Akhmetzhanov, Serial interval of novel coronavirus (COVID-19) infections. Int. journal infectious diseases (2020).

23. SW Park, et al., Forward-looking serial intervals correctly link epidemic growth to reproduction numbers. Proc. Natl. Acad. Sci. 118 (2021).

24. NG Davies, et al., Estimated transmissibility and impact of sars-cov-2 lineage b.1.1.7 in england. Science 372 (2021).

25. L Alvarez, Comparative analysis of the first wave of the COVID-19 pandemic in South Korea, Italy, Spain, France, Germany, the United Kingdom, the USA and the New-York state. MedRxiv (2020).

